# The physical and mental health benefits of touch interventions: A comparative systematic review and multivariate meta-analysis

**DOI:** 10.1101/2023.06.20.23291651

**Authors:** Julian Packheiser, Helena Hartmann, Kelly Fredriksen, Valeria Gazzola, Christian Keysers, Frédéric Michon

## Abstract

Introduction: Receiving touch is of critical importance for human well-being. A number of studies have shown that touch promotes mental and physical health. However, effect sizes differ considerably across studies and potential moderators of touch interventions remain unknown to this day.

**Methods:** We conducted a preregistered (CRD42022304281) systematic review and a large-scale multivariate multilevel meta-analysis encompassing 137 studies in healthy participants and patients (166 cohorts, 9617 participants and 643 effect sizes) in the meta-analysis and 75 additional studies as part of the systematic review to identify critical factors moderating touch intervention efficacy. Included studies always featured a touch vs. no touch control intervention with health outcomes as dependent variables.

**Results:** We found comparable and medium-sized (Hedges’ *g* ∼ 0.5) effects of touch on both mental and physical health. Touch interventions were especially effective in regulating cortisol levels (0.78 [0.24;1.31]) and increasing weight (0.65 [0.37;0.94]) in newborns, as well as in reducing pain (0.69 [0.48;0.89]), feelings of depression (0.59 [0.40;0.78]) and state (0.64 [0.44;0.84]) or trait anxiety (0.59 [0.40;0.77]) for adults and children. Comparing touch interventions involving objects or robots with humans resulted in similar physical (0.56 [0.24;0.88] vs. 0.51 [0.38;0.64]) but lower mental health benefits (0.34 [0.19;0.49] vs. 0.58 [0.43;0.73]). Adult clinical cohorts profited stronger in mental health domains compared to healthy individuals (0.63 [0.46;0.80] vs. 0.37 [0.20;0.55]) but showed comparable physical health benefits (0.53 [0.38;0.69] vs. 0.47 [0.29;0.65]). We found no difference in children and adults comparing touch applied by a familiar person or a health professional (0.51 [0.29;0.73] vs. 0.50 [0.38;0.61]) but parental touch was more beneficial in newborns (0.69 [0.50;0.88] vs. 0.39 [0.18;0.61]). Intervention frequency positively correlated with increased health benefits in adults and children while session duration did not show significant effects.

**Discussion:** Leveraging those factors that influence touch intervention efficacy will help maximize the benefits of future touch interventions and focus research in this field.

## 1. Introduction

The recent COVID-19 crisis has raised our awareness for the need to better understand the effects touch - and its reduction during social distancing - can have on our mental and physical well-being. Touch interventions, for example massages or kangaroo care, have been shown to have a wide range of both mental and physical health benefits over the lifespan, from facilitating growth and development to buffering against anxiety and stress in humans and animals alike (Ardiel & Rankin, 2010). Despite the substantial weight this literature gives to support the benefits of touch, it is also characterized by an immense variability in, for example, studied cohorts, type and duration of applied touch, measured health outcomes, and who actually applies the touch. While previous meta-analyses on this topic exist (e.g., Moyer et al., 2004), meaningful moderators could not yet be identified. However, understanding these variables is critical to tailor touch interventions, aiming to promote well-being both in healthy and clinical cohorts.

Here, we performed a pre-registered, large-scale systematic review and multi-level, multivariate meta-analysis to address this need with quantitative evidence for (a) the effect of touch interventions on physical and mental health and (b) which moderators influence the efficacy of the intervention. In particular, we ask whether and how strongly health outcomes depend on dynamics of the touching dyad (e.g., humans or robots/objects, familiarity, touch directionality), demographics (e.g., clinical status, age or sex), delivery means (e.g., type of touch intervention or touched body part) and procedure (e.g., duration or number of sessions). We did so separately for newborns and for children and adults, as the health outcomes in newborns differed substantially from those in the other age groups.

## 2. Materials and Methods

### Open science practices

All data and code are accessible in the corresponding <u>OSF project</u>. The systematic review was registered on PROSPERO (<u>CRD42022304281</u>) prior to the start of data collection. All deviations from the pre-registered plan can be found in the supplementary material.

### Inclusion and exclusion criteria

To be included in the systematic review, studies had to investigate the relationship between at least one health outcome (physical and/or mental) and a touch intervention, include explicit physical touch by another human, animal, or object as part of an intervention, and include an experimental and control condition/group that are differentiated by touch alone. The meta-analysis additionally required a between-subjects design (in order to clearly distinguish touch from no-touch effects). Studies that explicitly did not apply a randomized protocol were excluded prior to further analysis to reduce risk of bias (see Supplementary Methods for details).

### Data collection

We used Google Scholar, PubMed and Web of Science for our literature search, with no limitations regarding the publication date and using pre-specified search queries (see Figure 1 for an overview and the Supplementary Methods for the exact keywords used). Articles were assessed in French, Dutch, German or English. The databases were searched from 2^nd^ of December 2021 until the 01^th^ of October 2022. Two independent coders checked each paper against our inclusion and exclusion criteria. Inconsistencies between coders were checked and resolved by JP and HH. Studies excluded/included for the review and meta-analysis can be found on the <u>OSF project</u>.

**Figure 1.**
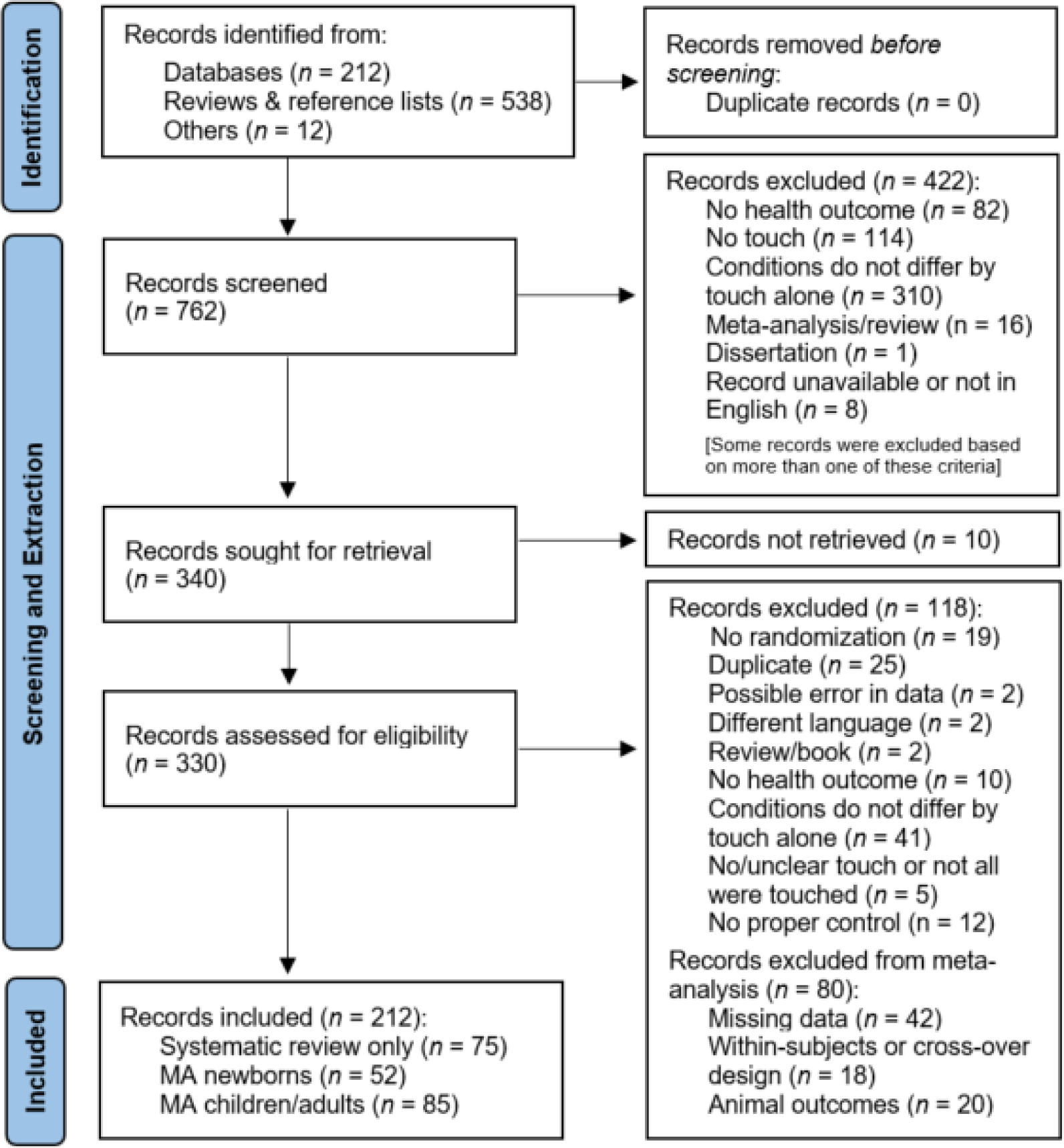
PRISMA 2020 flowchart detailing the identification and screening of identified records for the systematic review and meta-analysis <u>(Page et al., 2021)</u>. MA = Meta-analysis. Animal outcomes refer to outcomes measured in non-human species which were solely considered as part of a systematic review.

### Data extraction and preparation

Details for the data extraction process can be found in the supplementary material. From this data, Hedges’ *g* and its variance could be derived. Effect sizes were always computed between the experimental and the control group. Data extraction began on the 10^th^ of October 2022 and was concluded on the 25^th^ of February 2023. JP and HH oversaw the data collection process, and checked and resolved all inconsistencies between coders.

Health benefits of touch were always coded by positive summary effects whereas adverse health effects of touch were represented by negative summary effects. If multiple time points were measured for the same outcome on the same day after a single touch intervention, we extracted the peak effect size (in either the positive or negative direction). If the touch intervention occurred multiple times and health outcomes were assessed for each time point, we extracted data points separately. Measurements assessing long-term effects without explicit touch sessions in the breaks were excluded. In the case of multiple control groups, we always contrasted the touch group to the group that most closely matched the touch condition (e.g., relaxation therapy was preferred over standard medical care). A detailed data extraction guideline can be found on OSF. We extracted information from all moderators listed in the pre-registration (Table 1). A list of included and excluded health outcomes is presented in Supplementary Table S1. Authors of studies with possible effects but missing information to calculate those effects were contacted via email and asked to provide the missing data (response rate = 35.7%).

**Table 1.** Moderator description and coding specificities. Asterisks indicate that the moderator analyses can be found in the supplementary results.

| Moderator | Description | (Re)-Coding of variables |
| --- | --- | --- |
| **Health outcome**<br>(categorical) | On a broader scale, we used a dichotomous classification system (mental or physical health outcomes). To provide more detailed insight, we extracted health outcomes specifically as well. In newborns, mental health-like outcomes were rarely described as they are difficult to determine. These outcomes were assessed through behavioral means such as crying or parental assessment of mood. | All health outcomes were either classified as “Mental Health” (e.g., state anxiety, depression, or positive affect) or “Physical Health” outcomes (e.g., cortisol, pain or respiration). As specific health outcomes were highly diverse, we also re-coded them to form larger sub-groups. For example, back pain, stretch pain or headache were subsumed in a larger category of “pain”. Anxiety was divided into state and trait anxiety depending if the study inquired about momentary/state anxiety (e.g., through the STAI-S (Spielberger, 2012)) or longer-lasting symptoms of anxiety (at least over the past seven days through for example the STAI-T). |
| **Touch dyad**<br>(categorical) | Touch dyads were defined as human-human touch (humans being touched by other humans), human-robot touch (humans being touched by robots), human-object touch (humans touching objects) and human-animal touch (humans touch animals). This moderator was not investigated in the newborns meta-analysis since no study used object/robot/animal touch. | For children/adults, we re-coded this variable into “*Human-Human*” (all studies involving humans as toucher and touched individual) and “*Human-Object*” (all studies involving robots or objects touching or being touched by humans). Only a single study measured the effects of animal touch. We therefore excluded this study from moderator analysis in this category. |
| **Skin-to-skin contact**<br>(categorical) | For all studies, we extracted if the touch application involved skin-to-skin contact between the touching dyad. This moderator was not analyzed in the newborn meta-analysis as there was only a single study without skin-to-skin contact. | Studies were coded as skin-to-skin contact if the head, neck or hands were touched or if massage oils were used. By default, studies involving human-robot or human-object touch did not use skin-to-skin contact. |
| **Type of touch**<br>(categorical) | We assessed the type of touch that was applied in a given situation. The most prevalent type of touch was massage therapy for adults or kangaroo care in case of newborns. Other types of touch included for example hugs, gentle touch, tactile-kinesthetic stimulation or hand-holding. | Since massage therapy was the prime interest of previous studies as a touch intervention, we were interested in whether massages provide stronger benefits compared to other forms of touch. Since other touch forms were highly diverse and thus rarely found across multiple studies for children/adults, we re-coded this moderator dichotomously in “*Massage Therapy*” and “*Other Touch Types*”. Since a large number of studies |
|  |  | used “*Kangaroo Care*” in newborns, we included this type of touch as an additional factor level in the newborns meta-analysis. |
| **Clinical status**<br>(categorical) | Cohorts were both dichotomously divided into clinical and healthy individuals or the underlying disorder was specified. | Specific disorder types were re-coded into “*Cancer*” patients (e.g., breast cancer or bone cancer), patients with “*Neurological Disorders*” (e.g., Parkinson or dementia), patients with “*Pain Disorders*” (e.g., chronic back pain or fibromyalgia), patients with “*Depression Disorders*” (e.g., postpartum depression or major depressive disorder) or patients undergoing “*Surgery*” (e.g., aortic surgery or hip replacement). Patients with other kinds of disorders were subsumed in the “*Other Conditions*” category. For newborns, no re-classification was performed as the only clinical condition comprised premature birth with the exception of a single study investigating neonatal jaundice (C.-H. Lin et al., 2015). |
| **Touched body part\***<br>(categorical) | We recorded the body part (arm, leg, head or back) that was touched for moderation. | If more than one region was touched during the intervention, we classified it as “multiple regions”. This moderator was omitted for the newborns meta-analysis as all studies used multi-regional touch. |
| **Familiarity of the touch dyad**<br>(categorical) | Familiarity of the dyad was coded dichotomously as familiar or unfamiliar. | In case spouses, friends or parents were touching each other, we recorded the intervention as familiar touch. Otherwise, for example if touch was applied by the experimenter or health care personnel, the moderator was classified as unfamiliar. |
| **Familiarity of the location\***<br>(categorical) | Familiarity of the location was coded dichotomously as familiar or unfamiliar. | Interventions conducted in laboratories, hospitals or massage studios were classified as unfamiliar. Interventions at the participants’ homes were classified as familiar. For newborns, we omitted this moderator as there were no truly familiar locations in that case |
| **Sex of touched individual**<br>(continuous) | To account for potential sex differences, we assessed the sex of the person receiving the touch in a given cohort. | Since only seven effects were found for male child/adult cohorts and a large number of studies used mixed cohorts, we decided to convert this moderator from categorical to a continuous one using the ratio between sampled women/girls and men/boys. If a cohort comprised only women, the ratio was set to 100 whereas it was set to 0 if only men were sampled. |
| **Number of sessions**<br>(continuous) | We assessed how often the touch intervention occurred prior to measurement of the outcome. | Number of sessions was measured as a simple frequency of the touch interventions. |
| **Duration of touch**<br>(continuous) | We determined how long the touch was applied per session. | Duration was measured in minutes. Total duration (session duration * session number) was not investigated as a moderator as it was very strongly correlated with session number. |
| **Mean age**<br>(continuous) | Mean age was used as predictor of potential age differences. | Mean age was computed across the experimental and control group for a given effect. Since age did not vary in newborns, this moderator was excluded from the newborns meta-analysis. |
| **Directionality\*** | Studies were either coded as unidirectional or bi-directional touch. | Studies were classified as unidirectional whenever one person was a clear receiver and one person was a clear provider of touch. Studies were classified as bi-directional if touch was applied and received by both individuals in the dyad. This moderator was not investigated in newborns since touch interventions are never truly bi-directional in this case. If the health outcome was measured in the parent during kangaroo care, it was coded as bi-directional. |
| **Study location\***<br>(categorical) | We documented the region in which a study was conducted. | Study location was coded into North America, South America, Asia, Europe, Africa or Oceania. If a study explicitly mentioned for example a university or hospital as the location of study, we used this measure. In case there was no mention, the affiliation of the first author of the study was used instead. |

### Statistical analysis and risk of bias assessment

One meta-analysis was performed for adults, adolescents and children as outcomes were highly comparable. We will refer to this meta-analysis as the adult meta-analysis as children/adolescent cohorts were only targeted in a minority of studies. A separate meta-analysis was performed for newborns, as their health outcomes differed substantially from any other age group.

Data was analyzed using R (version 4.2.2) using the rma.mv function from the metafor package (Viechtbauer, 2010) in a multistep, multivariate and multilevel fashion (for details of random effects structure of the models, see Supplementary Analysis). We first looked at our primary moderators (mental vs. physical health) and how the effect sizes systematically varied as a function of our secondary moderators (e.g. human-human or human-object touch, duration, skin-to-skin presence, etc.).

Heterogeneity in the present study was assessed using Cochran’s *Q*, which determines whether the extracted effect sizes estimate a common population effect size. To assess small study bias, we visually inspected the funnel plot and used the variance as a moderator in the overarching meta-analyses. In the next step, we separately investigated all previously outlined moderators one-by-one. We also explored interaction effects of the primary moderators (mental and physical health) with all secondary moderators. Moderators were only investigated if sufficient power for the group analysis was present (see Supplementary Methods, Supplementary Figures S3 and S4, and Supplementary Table S1).

Post hoc tests were performed comparing mental and physical health benefits within each interacting moderator (e.g., mental vs. physical health benefits in cancer patients) and mental or physical health benefits across levels of the interacting moderator (e.g., mental health benefits in cancer vs. pain patients). Post hoc tests were uncorrected due to the pre-registered nature of the moderators. Data was visualized using forest plots and orchard plots (Nakagawa et al., 2021) for categorical moderators and scatter plots for continuous moderators.

Risk of bias was assessed for all studies included in both meta-analyses and the systematic review. A detailed description of assessed biases is outlined in the Supplementary Methods and an overview of the results is presented in Supplementary Figure S1).

## Results

### Touch interventions have a medium-sized effect on health for all ages

For adults, a total of *n* = 2841 and *n* = 2556 individuals in the touch and control groups, respectively, across 85 studies and 103 cohorts were included. The effect of touch overall was medium-sized (*g* = 0.52, Figure 2A). For newborns, we could include 63 cohorts across 52 studies comprising a total of *n* = 2134 and *n* = 2086 newborns in the touch and control groups, respectively, with an overall effect almost identical to the older age group (*g* = 0.56, Figure 2B) suggesting that, despite distinct health outcomes, touch interventions show comparable effects across newborns and adults. Sufficient power to detect such effect sizes was rare in individual studies (see Supplementary Figure S2 and S3). No individual effect size from either meta-analysis was overly influential (Cook’s *D* < 0.06). The benefits were similar for mental and physical outcomes (mental vs physical, adults: *p* = 0.432, Figure 2C, newborns: *p* = 0.284, Figure 2D). Results for studies that were only part of the systematic review mimicked the result pattern of the overall meta-analysis and can be found in the Supplementary Results (Section “Systematic Review”).

**Figure 2.**
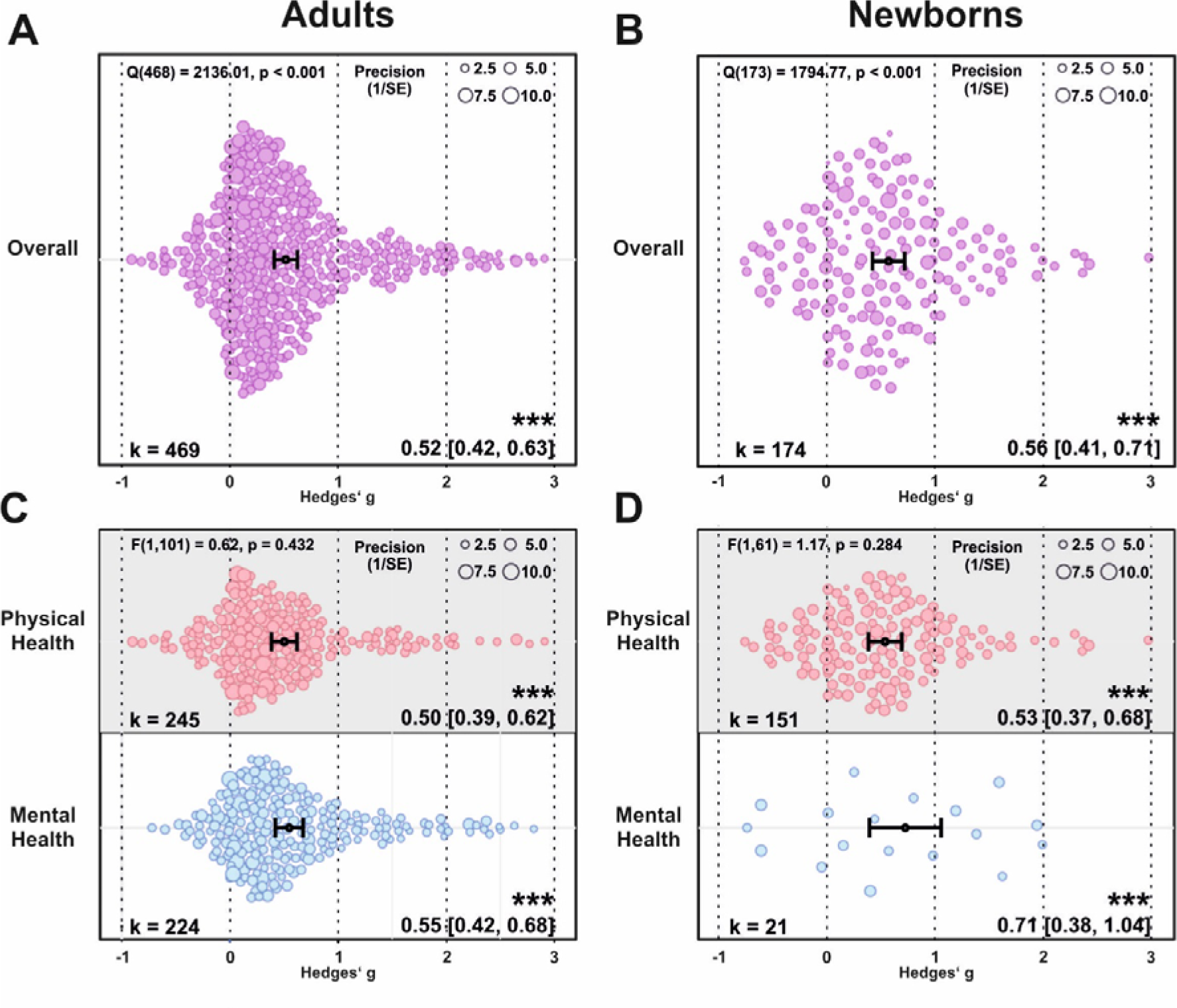
Benefits of touch on physical and mental health. (A) Orchard plot illustrating the overall benefits across all health outcomes for adults/children. (B) same as A for newborns. (C-D) same as (A-B) but separating the results for physical vs mental health benefits. Each dot reflects a measured effect and the number of effects (k) included in the analysis is depicted in the bottom left. Mean effects and 95% CIs are presented in the bottom right, the heterogeneity Q statistic is presented in the top left. Asterisks indicate the overall effect being significant from a null effect (*** p < .001, ** p < .01, * p < .05). Dot size reflects precision of each individual effect (larger = higher precision). Small study bias for the overall effect in the adult meta-analysis was significant (p < .001, see Figure S6) but did not reach significance for the newborn meta-analysis (p = .070, see Figure S7).

Based on the overall effect of both meta-analyses as well as their median sample sizes, the minimum number of studies necessary for subgroup analyses to achieve 80% power was *k* = 9 effects for adults and *k* = 8 effects for newborns (see Supplementary Figures S4 and S5). Assessing specific health outcomes with sufficient power in more detail in adults (Figure 3A) revealed smaller benefits to sleep and heart rate parameters, moderate benefits to positive and negative affect, diastolic blood and systolic blood pressure, mobility and reductions of the stress hormone cortisol, and larger benefits to trait and state anxiety, depression, fatigue and pain. Post hoc tests revealed stronger benefits for pain, state anxiety, depression and trait anxiety compared to respiratory, sleep and heart rate parameters (all *p*s < .05). Reductions in pain and state anxiety were increased compared to reductions in negative affect (both *p*s < .05). Benefits to pain symptoms were higher compared to benefits to positive affect (*p* = .030). Finally, touch resulted in larger benefits to cortisol release compared to heart rate and respiratory parameters (both *p*s < .05).

**Figure 3.**
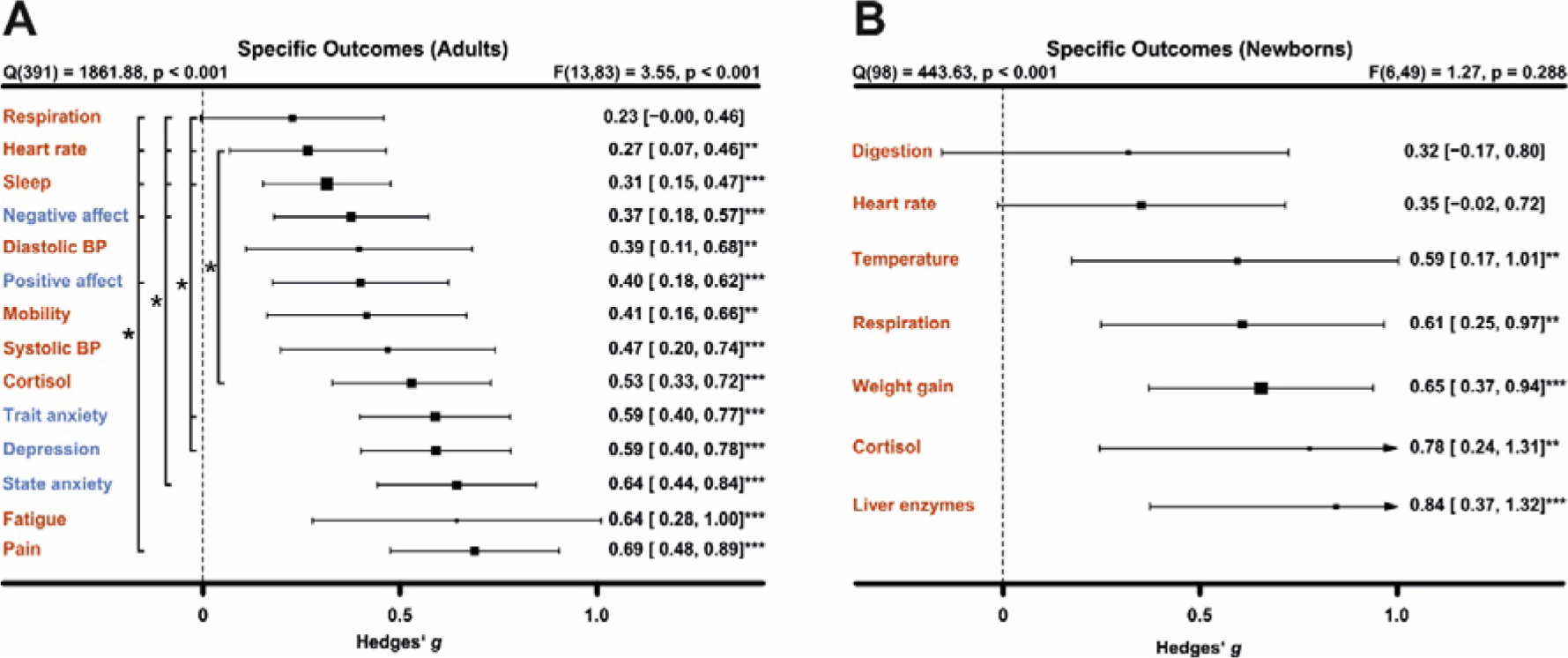
Forest plot for all specific health outcomes with sufficient effects to warrant further analysis. The type of health outcomes measured differed between adults (A) and newborns (B) and were thus analyzed separately. Numbers on the right represent the mean effect, its 95% CI in square brackets and the significance level estimating the likelihood that the effect is equal to zero. The F-value in the top right represents a test of the hypothesis that all effects within the subpanel are equal. The Q statistic represents heterogeneity. Asterisks indicate the overall effect being significant from a null effect (*** p < .001, ** p < .01, * p < .05). Physical outcomes are marked in red, mental outcomes are marked in blue.

In newborns, only physical health effects had sufficient data for further analysis. We found no benefits for digestion and heart rate parameters. All other health outcomes (cortisol, liver enzymes, respiration, temperature regulation and weight gain showed medium to large effects (Figure 3B). We found no significant differences among any specific health outcomes.

### Non-human touch and skin-to-skin contact

In some situations, a fellow human is not readily available to provide affective touch, raising the question of the efficacy of touch delivered by objects and robots (Eckstein et al., 2020). Overall, we found humans engaging in touch with other humans or objects to have medium-sized health benefits in adults, without significant differences (*p* = .295, Figure 4A). However, differentiating physical vs mental health benefits revealed similar benefits for human and object touch on physical health outcomes, but larger benefits on mental outcomes when humans were touched by humans (*p* = .022, Figure 4B). It needs to be noted that object-touch still showed a significant effect (see also Supplementary Figure S8 for the corresponding Orchard plot).

**Figure 4.**
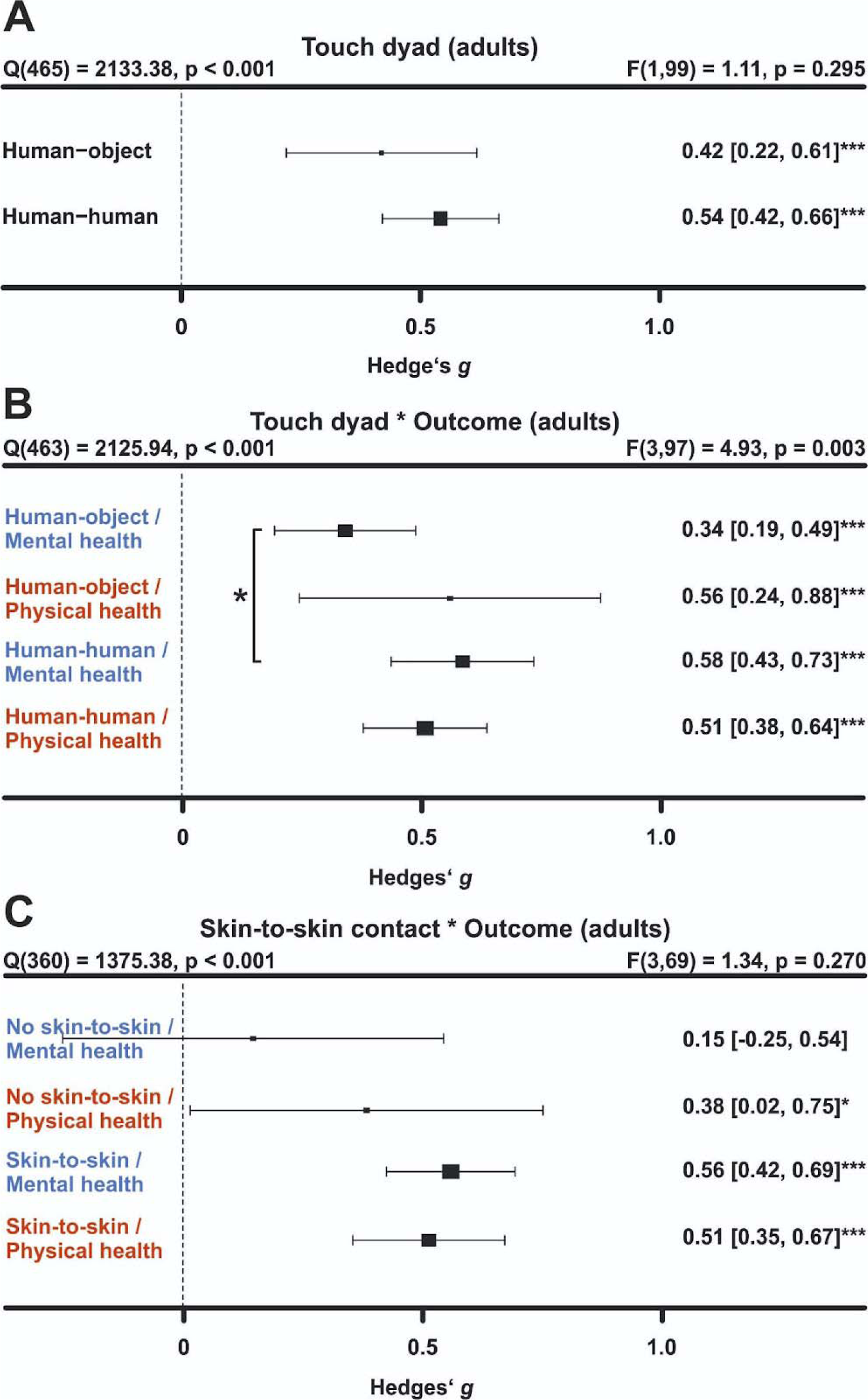
Influence of the touching dyad in adults. (A) Forest plot comparing humans vs objects touching a human on health outcomes overall. (B) same as A, but separately for mental vs physical health outcomes. In C), we removed all object studies to identify if missing skin-to-skin contact is the relevant mediator of higher mental health effects in human-human interactions. Numbers on the right represent the mean effect, its 95% CI in square brackets and the significance level estimating the likelihood that the effect is equal to zero. The F-value in the top right represents a test of the hypothesis that all effects within the subpanel are equal. The Q statistic represents heterogeneity. Asterisks indicate the overall effect being significant from a null effect (*** p < .001, ** p < .01, * p < .05). Physical outcomes are marked in red, mental outcomes are marked in blue.

We considered the possibility that this effect is due to missing skin-to-skin contact in human-object interactions. Thus, we investigated human-human interactions with and without skin-to-skin contact (Figure 4C). In line with the hypothesis that skin-to-skin contact is highly relevant, we again found stronger mental health benefits in presence of skin-to-skin contact that however did not achieve nominal significance (*p* = .055), likely because skin-to-skin contact was rarely absent in human-human interactions leading to a decrease in power for the analysis. Results for skin-to-skin contact as moderator can be found in Supplementary Figure S9.

### Influences of type of touch

The large majority of touch interventions comprise massage therapy in adults and kangaroo care in newborns. However, comparing the different types of touch explored across studies did not reveal significant differences in effect sizes based on touch-type, be it on overall health benefits (adults, *p* = .916; newborns *p* = .361, Figure 5A,B) or comparing different forms of touch separately for physical or for mental health benefits (pairwise comparisons *p* > .325, Figure 5C/D; see also Supplementary Figure S10/11 for the corresponding Orchard plots). This suggests that touch types may be flexibly adapted to the setting of every touch intervention.

**Figure 5.**
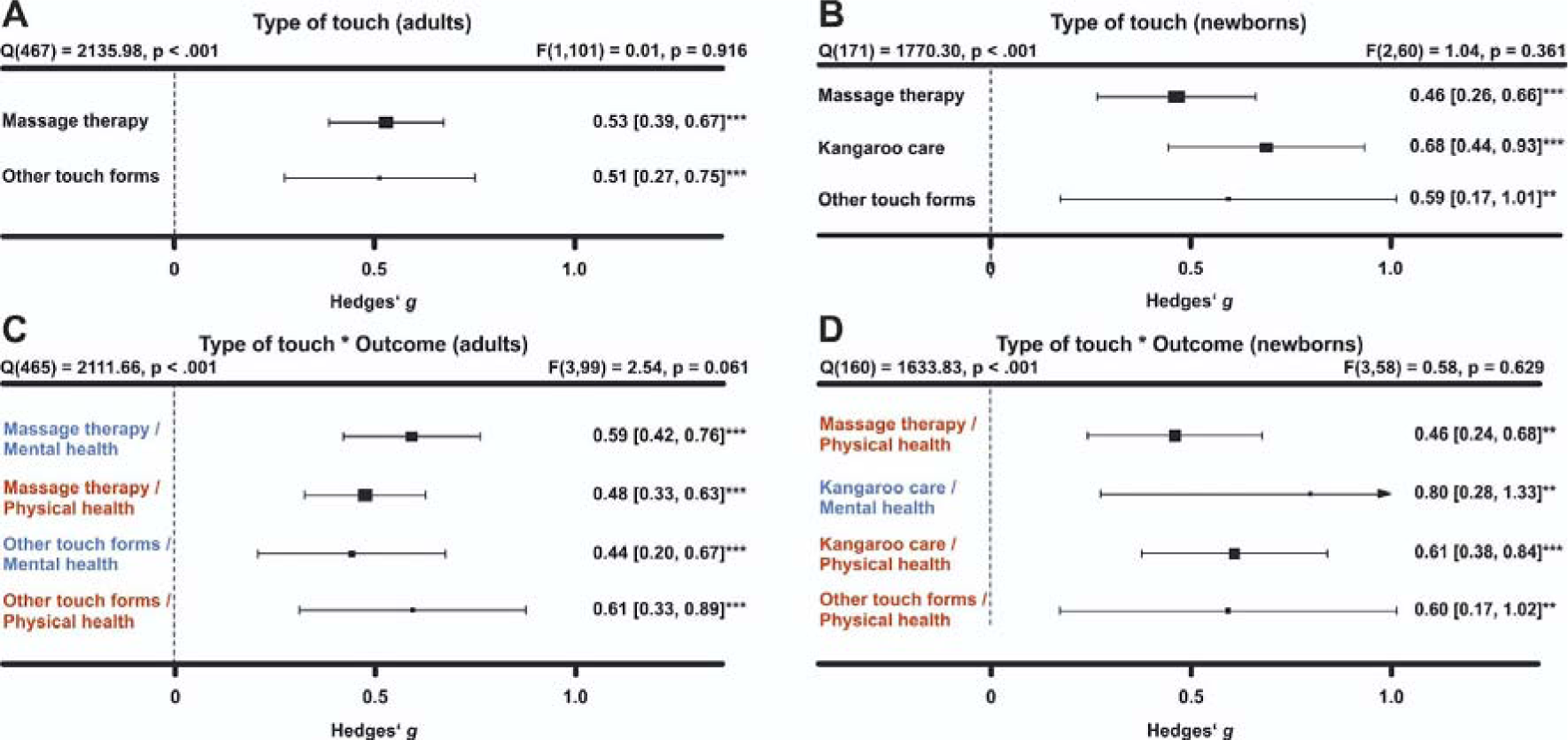
Effect of type of touch: (A) Forest plot of health benefits comparing massage therapy vs. other forms of touch in adult cohorts. (B) Forest plot of health benefits comparing massage therapy, kangaroo care and other forms of touch for newborns. (C) same as (A) separating mental and physical health benefits. (D) same as B but separating mental and physical health outcomes where possible. Note that an insufficient number of studies assessed mental health benefits of massage therapy or other forms of touch to be included. Numbers on the right represent the mean effect, its 95% CI in square brackets and the significance level estimating the likelihood that the effect is equal to zero. The F-value in the top right represents a test of the hypothesis that all effects within the subpanel are equal. The Q statistic represents heterogeneity. Asterisks indicate the overall effect being significant from a null effect (*** p < .001, ** p < .01, * p < .05). Physical outcomes are marked in red, mental outcomes are marked in blue.

### The role of clinical status

Most research on touch interventions has focused on clinical samples, but are benefits restricted to clinical cohorts? We found health benefits to be significant in clinical and healthy populations (Figure 6), whether all outcomes are considered (Figure 6A,B), or physical and mental outcomes are separated (Figure 6C/D, see Supplementary Figure S12/13 for the corresponding Orchard plots). In adults, however, we found higher mental health benefits for clinical populations compared to healthy ones (Figure 7C; *p* = .037).

**Figure 6.**
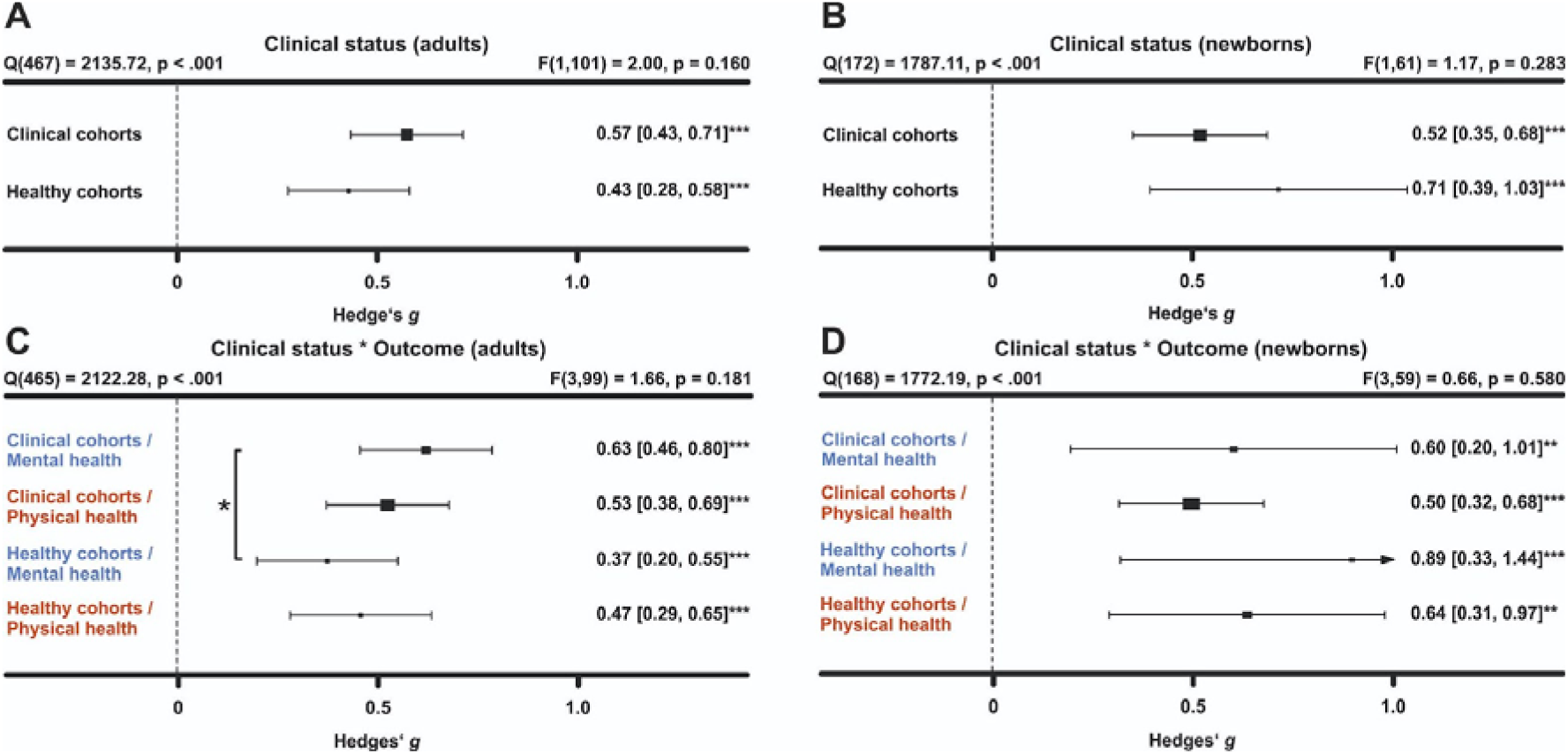
Comparing health benefits for clinical vs. healthy cohorts. (A) health benefits for clinical cohorts of adults vs. healthy cohorts of adults. (B) same as A for newborn cohorts. (C) same as A, but separating mental vs physical health benefits. (D) same as B but separating mental vs physical health benefits. Numbers on the right represent the mean effect, its 95% CI in square brackets and the significance level estimating the likelihood that the effect is equal to zero. The F-value in the top right represents a test of the hypothesis that all effects within the subpanel are equal. The Q statistic represents heterogeneity. Asterisks indicate the overall effect being significant from a null effect (*** p < .001, ** p < .01, * p < .05). Physical outcomes are marked in red, mental outcomes are marked in blue.

**Figure 7.**
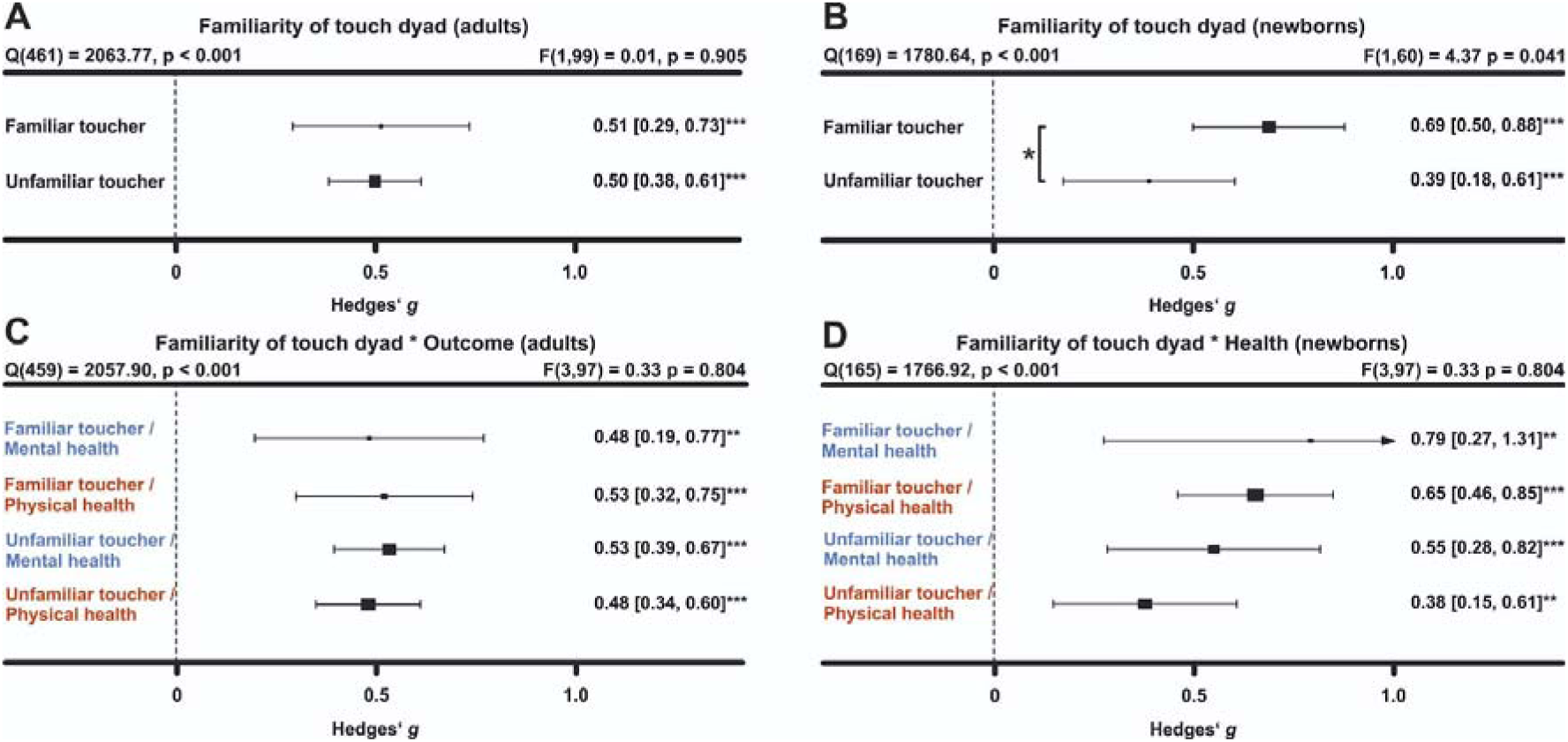
Comparing health benefits for familiar vs unfamiliar touchers. (A) health benefits for being touched by a familiar (e.g. partners, family members or friends) vs unfamiliar toucher (health professional). (B) same as A for newborn cohorts. (C) same as A, but separating mental vs physical health benefits. (D) same as B but separating mental vs physical health benefits. Numbers on the right represent the mean effect, its 95% CI in square brackets and the significance level estimating the likelihood that the effect is equal to zero. The F-value in the top right represents a test of the hypothesis that all effects within the subpanel are equal. The Q statistic represents heterogeneity. Asterisks indicate the overall effect being significant from a null effect (*** p < .001, ** p < .01, * p < .05). Physical outcomes are marked in red, mental outcomes are marked in blue.

A more detailed analysis of specific clinical conditions revealed positive mental and physical health benefits for almost all assessed clinical disorders. Differences between disorders were not found with the exception of increased effectiveness of touch interventions in neurological disorders (see Supplementary Figure S14).

### Familiarity of the touching dyad

Touch interventions can either be performed by familiar touchers (partners, family members or friends) or by unfamiliar touchers (health care professionals). In adults, we did not find an impact of familiarity of the toucher (*p* = .905; see Figure 7 and Supplementary Figure S15/16 for the corresponding Orchard plots). Similarly, investigating the impact on mental and physical health benefits specifically, no significant differences could be detected suggesting that familiarity is irrelevant in adults.

In contrast, touch applied by the parents, almost exclusively the mother, was significantly more beneficial compared to unfamiliar touch (*p* = .041) for newborns. Investigating mental and physical health benefits specifically revealed no significant differences.

### Frequency and duration of touch interventions

How often and for how long should touch be delivered? For adults, the median touch duration across studies was 20 minutes and the median number of touch interventions was four sessions with an average time interval of 2.3 days in between each session. For newborns, the median touch duration across studies was 17.5 minutes and the median number of touch interventions was seven sessions with an average time interval of 1.3 days in between each session.

Delivering more touch sessions increased benefits in adults, whether overall, physical or mental benefits were measured (all *p*s < .008, Figure 8A). A closer look at specific outcomes for which sufficient data was available revealed that positive associations between the number of sessions and outcomes were found for trait anxiety, depression and pain (*all ps* < .001), indicating a need for repeated sessions to improve these adverse health outcomes. Neither increasing the number of sessions for newborns, nor increasing the duration of touch per session in adults or newborns increased health benefits, be they physical or mental (Figure 8, B-D). For continuous moderators in adults, we also looked at specific health outcomes as sufficient data was generally available for further analysis. Surprisingly, we found significant negative associations between touch duration and reductions of cortisol (*p* = .012) and heart rate parameters (*p* = .029).

**Figure 8.**
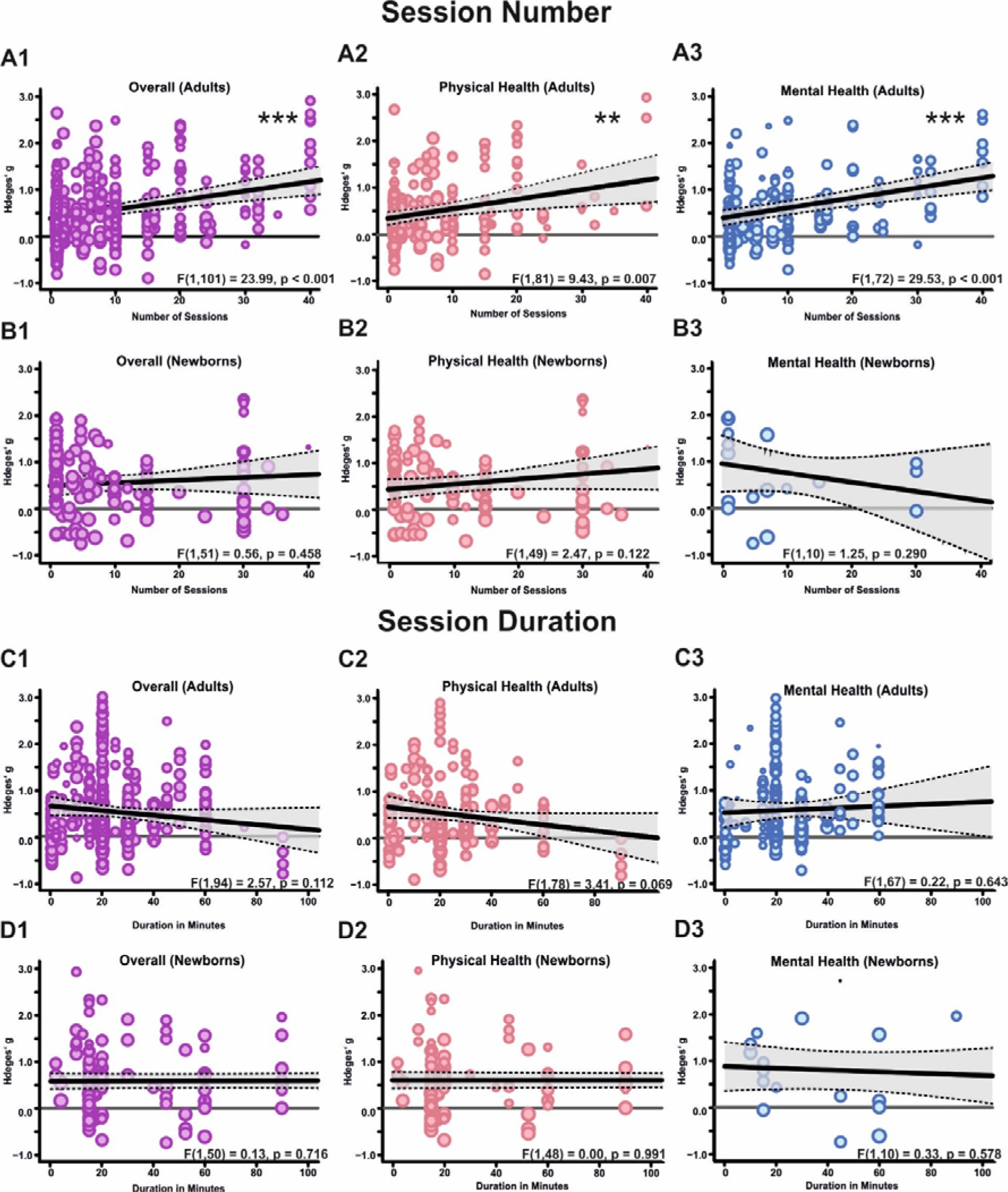
Effect of the number of sessions and their duration on health outcomes. (A) Meta-regression analysis examining the association between the number of sessions applied and the effect size in adults, either on overall health benefits (A1), or for physical (A2) or mental benefits (A3) separately. (B) same as A for newborns. (C,D) same as A,B but for the duration of the individual sessions. Each dot represents an effect size, its size indicates the precision of the study (larger = better). The asterisks in each panel represent the result of a regression analysis testing the hypothesis that the slope of the relationship is equal to zero. *** = p < 0.001. The shaded area around the regression line represents the 95% CI.

### Demographic influences of sex, age and regional background

We used the ratio between women and men in the sample as a proxy for sex-specific effects. Sex ratios were heavily skewed towards larger numbers of women in each cohort (median = 83% women) and we could not find significant associations between sex ratio and overall, mental or physical health benefits (all *ps* > .587). For specific outcomes that could be further analyzed, we found a significant positive association of sex ratio with reductions in cortisol secretion (*p* = .033) suggesting stronger benefits in women. In contrast to adults, sex ratios were balanced in newborns samples (median = 53% girls).There was no significant association with overall and physical health benefits of touch (all *ps* > .359). Mental health benefits did not provide sufficient data for further analysis.

The median age in the adult meta-analysis was 42.6 years (range: 4.5 - 88.4). There was no association between age and the overall, mental and physical health benefits of touch (all *ps* > .745). Looking at specific health outcomes, we found significant positive associations between mean age and improved positive affect (*p* = .030) as well as systolic blood pressure (*p* = .036).

## Discussion

The key aim of the present study was twofold: to provide an estimate of the effect size of touch interventions, and to disambiguate moderating factors to tailor future interventions more precisely. Overall, touch interventions were beneficial for both physical and mental health with a medium effect size. Our work illustrates that touch interventions are best suited for reducing pain, depression, and anxiety in adults and children, as well as for increasing weight gain in newborns. In general, both massages (Field, 2016; Moyer et al., 2004) and other types of touch, such as gentle touch, kangaroo care or acupressure, were equally beneficial.

While it seems to be less critical which touch intervention is applied, the frequency of interventions seem to matter. More sessions were positively associated with the improvement of trait outcomes such as depression and anxiety but also pain reductions in adults. In contrast to session number, increasing the duration of individual sessions did not improve health effects. In fact, we found some indications of negative relationships in adults for cortisol and blood pressure. This could be due to habituating effects of touch on the sympathetic nervous system and HPA-axis, ultimately resulting in diminished effects with longer exposure, or decreased pleasantness ratings of affective touch with increasing duration (Bendas et al., 2021). For newborns, we could not support previous notions that the duration of the touch intervention is linked to benefits in weight gain (Charpak et al., 2021). Thus, an ideal intervention protocol does not seem to have to be excessively long.

A critical issue highlighted in the pandemic is the lack of touch due to social restrictions (Packheiser et al., 2023). To accommodate the need for touch in individuals with small social networks (e.g., institutionalized or isolated individuals), touch interventions using objects/robots have been explored in the past (for review, see Eckstein et al., 2020). We show here that touch interactions outside of the human-human domain are beneficial for mental and physical health outcomes. Importantly, object/robot touch was not as effective in improving mental health as human-applied touch. A sub-analysis of missing skin-to-skin contact among humans indicated that mental health effects of touch might be mediated by the presence of skin-to-skin contact. Thus, it seems profitable to include skin-to-skin contact in future touch interventions in line with previous findings in newborns (Whitelaw et al., 1988). In robots, recent advancements in synthetic skin (Yogeswaran et al., 2015) should be investigated further in that regard.

Touch was beneficial for both healthy and clinical cohorts. This data is critical as most previous meta-analytic research has focused on individuals diagnosed with clinical disorders (citations). For mental health outcomes, we found larger effects in clinical cohorts. A possible reason could relate to increased touch wanting (Durkin et al., 2021) in patients: For example, loneliness often co-occurs with chronic illnesses (Rokach et al., 2006), which is linked to depressed mood and feelings of anxiety (Palgi et al., 2020). Touch can be used to counteract this negative development (Heatley-Tejada et al., 2020, Packheiser et al., 2022). In adults and children, knowing the toucher did not influence health benefits. In contrast, familiarity affected overall health benefits in newborns, with parental touch being more beneficial than touch applied by medical staff. Previous studies have suggested that early skin-to-skin contact and exposure to maternal odor is critical for a newborn’s ability to adapt to a new environment (Porter, 2007), supporting the notion that parental care is difficult to substitute in this time period. Research on sex differences in touch processing and benefits is sparse (but see Russo et al., 2020). Our results suggest that at least buffering effects against physiological stress are stronger in women. This is in line with increased buffering effects of hugs in women compared to men (Berretz et al., 2021). The female biased samples, however, beg for more research in men or non-binary individuals. Our data further suggests that increasing age was related to a higher benefit through touch, but only for cortisol and systolic blood pressure. These findings could potentially be attributed to higher basal cortisol levels (Seeman et al., 2001) and blood pressure (Hawkley et al., 2006) with increasing age, allowing for a stronger modulation of these parameters. Our results offer many promising avenues to improve future touch interventions, but they also need to be discussed in light of their limitations. For one, an important prerequisite for touch to be beneficial is its perceived pleasantness which could not be accounted for in our meta-analysis. The level of pleasantness associated with being touched is modulated by several parameters (Saarinen et al., 2021) including cultural acceptability (Burleson et al., 2019), perceived humanness (Wijaya et al., 2020), or a need for touch (Golaya, 2021), which could explain the observed differences for certain moderators, such as human-human vs. robot-human interaction. Moreover, the fact that secondary categorical moderators could not be investigated with respect to specific health outcomes, due to the lack of data points, limits the specificity of our conclusions. It thus remains unclear whether, for example, a decreased mental health benefit in the absence of skin-to-skin contact is linked mostly to decreased anxiolytic effects, changes in positive/negative affect or something else. Since these health outcomes are however highly correlated (Ng et al., 2019), it is likely that such effects are driven by multiple health outcomes. Finally, it needs to be noted that blinding towards the experimental condition is essentially impossible in touch interventions. Although we compared the touch intervention to other interventions, such as relaxation therapy, as control whenever possible, contributions of placebo effects cannot be ruled out.

In conclusion, we show clear evidence that touch interventions are beneficial across a large number of both physical and mental health outcomes, both for healthy and clinical cohorts, and for all ages. These benefits, while influenced in their magnitude by study cohorts and intervention characteristics, were robustly present promoting the conclusion that touch interventions can be systematically employed across the population to preserve and improve our health.

## Data Availability

All data are available under the following link: https://osf.io/c8rvw/?view_only=6298307ead2e423eb20f9675f843de0d

## Author contributions

**JP:** Conceptualization, Methodology, Formal analysis, Investigation, Data Curation, Writing - Original Draft, Writing - Review & Editing, Visualization, Supervision, Project administration. **HH:** Conceptualization, Methodology, Formal analysis, Investigation, Data Curation, Writing - Original Draft, Writing - Review & Editing, Visualization, Supervision, Project administration. **KF:** Investigation, Data Curation, Writing - Review & Editing. CK: Writing - Review & Editing, Conceptualization. **VG:** Writing - Review & Editing, Conceptualization. **FM:** Conceptualization, Methodology, Formal analysis, Investigation, Writing - Original Draft, Writing - Review & Editing.

## Acknowledgements

We thank Aline Frick and Eva Chris for supporting the initial literature search and coding. We also thank Aljoscha Dreisoerner, Tiffany Field, Sander Koole, Cynthia Kuhn, Maria Henricson, Laura Frey Law, Joy Fraser, Marianne Cumella Reddan, and Jacqui Stringer who kindly responded to our data requests and provided additional information or data with respect to single studies.

