## Supplementary Information for "The physical and mental health benefits of touch interventions: A comparative systematic review and multivariate meta-analysis"

^+^ Shared first authorship

***Supplementary Material***

*Supplementary Methods*

*Deviations from pre-registration*

**Deviation #1**: During our initial screening for the systematic review, we were confronted with a large number of potential health outcomes to look at. This observation of multivariate outcomes led us to register an amendment during data collection (but prior to any effect size or moderator screening). In doing so, we aimed to additionally extract meta-analytic effects for a more quantitative assessment of our review question that can account for multivariate data reporting and dependencies of effects within the same study. Furthermore, as we noted a severe lack of studies with respect to health outcomes for animals during the inclusion assessment for the systematic review, we decided that the meta-analysis would only focus on outcomes that could be meaningfully analyzed on the meta-analytic level and therefore only included health outcomes of human participants. Furthermore, since health outcomes differ significantly depending on the age of the source population, we report two separate meta-analyses, one for children/adults, and one for newborns. All deviations from the pre-registration are reported below. We had initially registered two separate meta-analyses for adults and children, but noticed during the data collection, that the separation made more sense for newborns due to similar health outcomes in children and adults, but vastly different health outcomes in newborns.

**Deviation #2**: In the pre-registration, we did not explicitly exclude non-randomized trials. Since an explicit use of non-randomization for group allocation significantly increases the risk of bias, we decided to exclude them a posteriori from data analysis.

**Deviation #3**: In the pre-registration, we also outlined a tertiary moderator, namely benefits of touch application vs. touch reception. Thís level was ignored since no included study specifically investigated the benefits of touch application by itself.

**Deviation #4**: In the pre-registration, we suggested using the RoBMA function (Maier et al., 2022) to provide a Bayesian framework that allows for a more accurate assessment of publication bias beyond small study bias. Unfortunately, neither multilevel nor multivariate data structures are supported by the RoBMA function. For that reason, we did not further pursue this analysis as the hierarchical nature of the data would not be accounted for.

**Deviation #5**: Beyond the pre-registered in- and exclusion criteria, we also excluded dissertations due to their lack of peer-review.

**Deviation #6**:  In the pre-registration, we stated to investigate the impact of sex of the person applying the touch. This moderator was not further analyzed as this information was rarely given and the touching individuals were almost exclusively women (adults/children: 7 males, 24 mixed, 85 females; newborns: 3 males, 17 mixed, 80 females).

**Deviation #7**: Time span of the touch intervention as assessed by subtracting the final day of the intervention from the first day was not investigated further due to its very high correlation with the number of sessions (*r*(461) = 0.81 in the adult meta-analysis, *r*(145) = 0.84 in the newborn meta-analysis).

*Search queries*

We used the following keywords to search the chosen databases. Agents (human vs. animal vs. object vs. robot) and touch outcome (physical vs. mental) were searched separately.

1. **TOUCH**: Touch AND Social OR Affective OR Contact OR Tactile interaction OR Hug OR Massage OR Embrace OR Kiss OR Cradling OR Stroking OR Haptic interaction OR tickling
2. **AGENT**: Object OR Robot OR human OR (animal OR rodent OR primate)
3. **MENTAL OUTCOME**: Health AND mood OR Depression OR Loneliness OR happiness OR life satisfaction OR Mental Disorder OR well-being OR welfare OR dementia OR psychological OR psychiatric OR anxiety OR Distress
4. **PHYSICAL OUTCOME**: Health AND Stress OR Pain OR cardiovascular health OR infection risk OR immune response OR blood pressure OR heart rate

*Data extraction*

The full study lists for excluded and included studies can be found [here](https://osf.io/c8rvw/?view_only=978a5dace61f4a43806b2d39895844d3) in the file “Study_lists_final.xlsx”. After finalizing the list of included studies for the systematic review, we added columns for moderators and the coding schema for our meta-analysis per our updated registration. Then, each study was assessed for its eligibility in the meta-analysis by two independent coders (JP, HH, KF or FM). To this end, all coders followed an a-priori specified procedure: First, the PDF was skimmed for possible effects to extract, and the study was excluded if no PDF was available or the study was in a language different than the ones specified in Data collection. Effects from studies that met the inclusion criteria were extracted from all studies listing descriptive values or statistical parameters to calculate effect sizes (means and standard deviations/standard errors/confidence intervals, sample sizes, *F*-values, *t*-values, *t*-test *p*-values, or frequencies). If only *p*-value thresholds were reported (e.g., *p* < .01), we used this, most conservative, value as the *p*-value to calculate the effect size (e.g., *p* = .01). If only the total sample size was given but that number was even and the participants were randomly assigned to each group, we assumed equal sample sizes for each group. If delta change scores (e.g. pre- to post-touch intervention) were reported, we used those over post-touch only scores. In case frequencies were 0 when frequency tables were used to determine effect sizes, we used a value of 0.5 as a substitute to calculate the effect (default setting in the metafor function; Viechtbauer, 2010).

*Risk of bias assessment and statistical analysis*

 In a first step, we assessed the risk of bias for the following parameters:

(1) **Bias from randomization** including whether a randomization procedure was performed, whether it a between- or within-participant design, and whether there were any baseline differences for demographic or dependent variables.
(2) **Sequence bias** resulting from a lack of counterbalancing in within-subject designs.
(3) **Performance bias** resulting from the participants or experiments not being blinded to the experimental conditions.
(4) **Attrition bias** resulting from different dropout rates between experimental groups.

The results can be found in Supplementary Figure S1. It should be noted that few studies in the adult meta-analysis did not explicitly mention randomization as part of their protocol. However, since these studies never showed any baseline differences in all relevant variables, we assumed that randomization was performed but not mentioned. An exclusion of these studies from the overall meta-analysis does not alter the main conclusions of the manuscript. Sequence bias was of no concern for studies for the meta-analysis since cross-over designs were excluded. It was however assessed for studies within the scope of the systematic review.

We calculated an overall effect of touch interventions across all studies, cohorts and health outcomes. To account for the hierarchical structure of the data, we used a multilevel structure with random effects at the study, cohort and effects level. Furthermore, we calculated the variance-covariance matrix of all data points to account for the dependencies of measured effects within each individual cohort and study. The variance-covariance matrix was calculated by default with an assumed correlation of effect sizes within each cohort of rho = 0.6. In addition to these procedures, we used robust-variance estimation with cluster-robust inference at the cohort level. This step is recommended to more accurately determine the confidence intervals in complex multivariate models (Pustejovsky & Tipton, 2022).

 To determine whether individual effects had a strong influence on our results, we calculated Cook’s distance (*D*). Here, a threshold of *D* > 0.5 was used to qualify a study as influential (Cook, 2011). Prior to any sub-group analysis, the overall effect size was used as input for power calculations. Power calculation for random-effects models further requires a sample size for each individual effect as well as an approximation of the expected heterogeneity between studies. For the sample size input, we used the median sample size in each of our studies. For heterogeneity, we assumed a value between medium and high levels of heterogeneity (*I*² = 62.5%; Higgins et al., 2003), as moderator analyses typically aim at reducing heterogeneity overall. Subgroups were only further investigated if the number of observed effects achieved ~80% power under these circumstances to allow for a more robust interpretation of the observed effects (see Supplementary Figures S4/5).

*Supplementary Results*

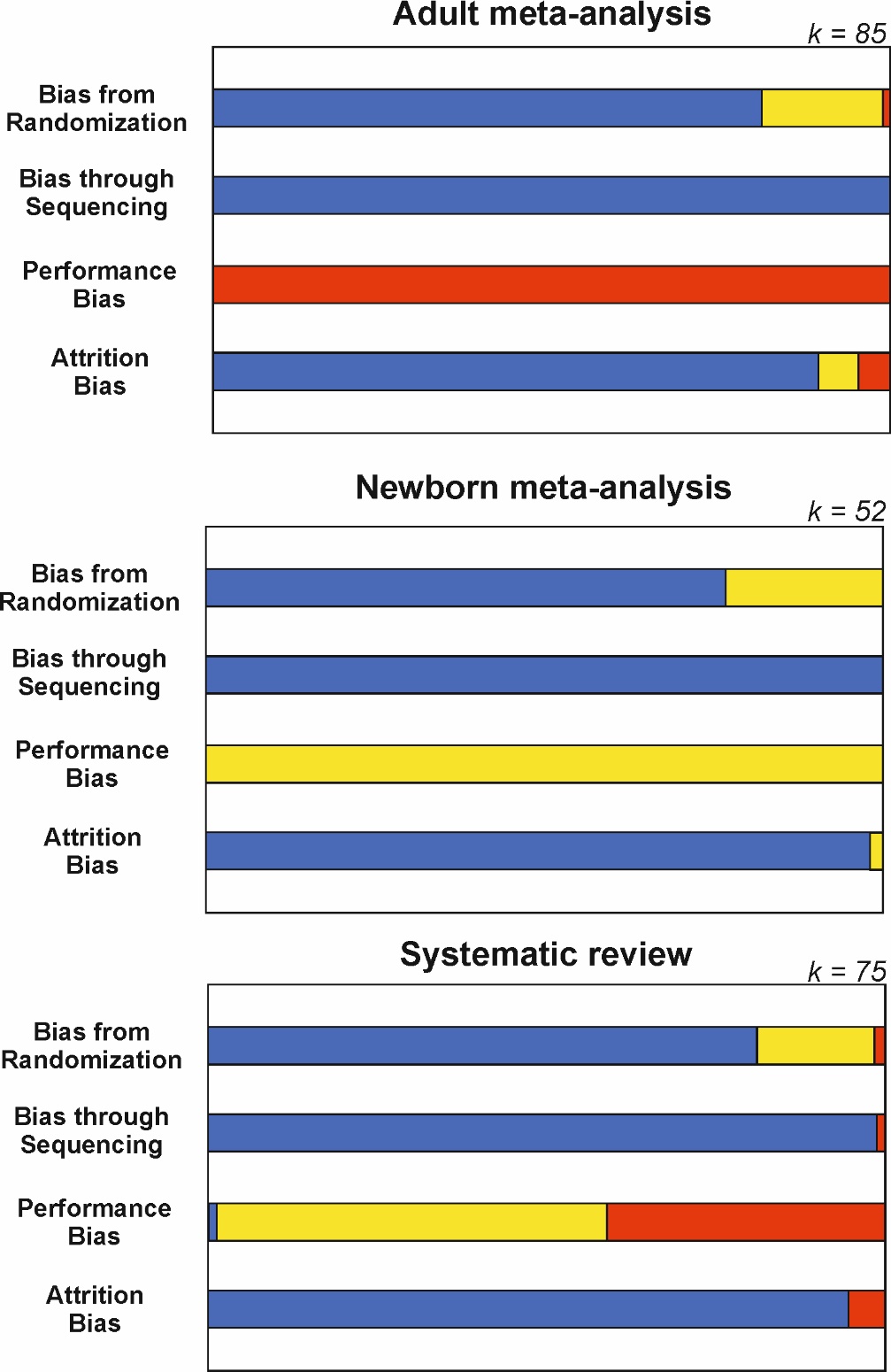

**Figure S1. Risk of bias overview.** Results from the assessment of randomization, sequence, performance and attrition bias. Not that sequence bias was only possible in within-subjects cross-over designs that were excluded from both meta-analyses. Furthermore, performance bias was always high in the adult meta-analysis as blinding of the participants and experimenters to the experimental conditions was not possible due to the nature of the intervention. For studies with newborns and animals, we assessed the performance bias as medium since neither newborns or animals were aware as being part of an experimental group. Blue = Low risk of bias, Yellow = Medium risk of bias, Red = High risk of bias. Bar length corresponds to percentages.

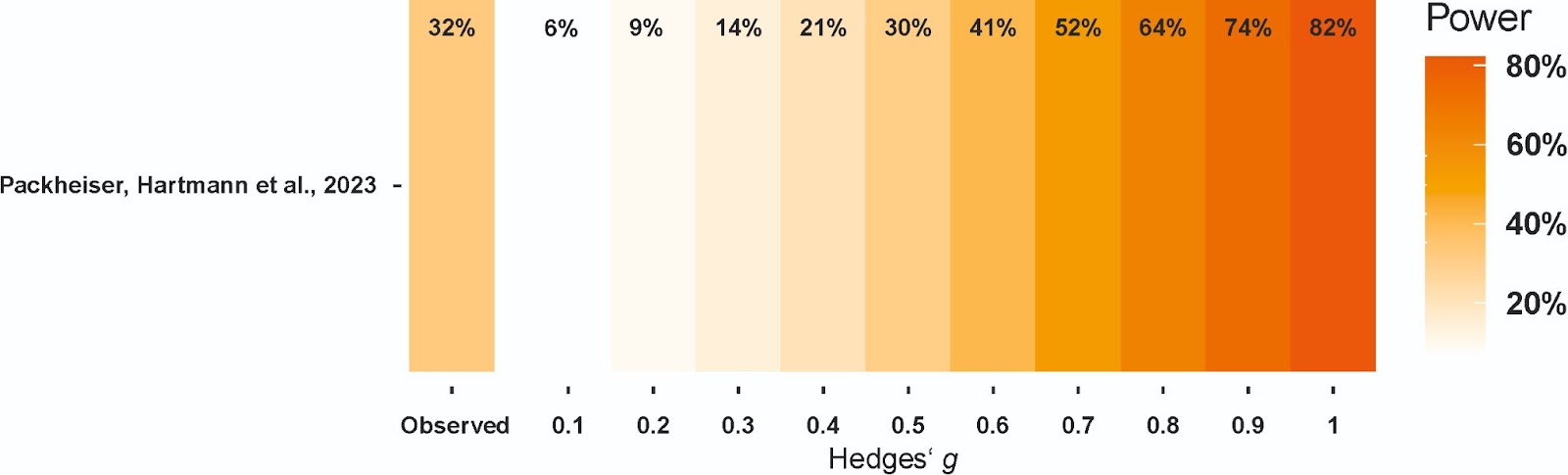

***Figure S2. Firepower plot (Quintana, 2022) for the meta-analysis of children/adults studies.*** *The median power of each individual study in this meta-analysis was 32% to detect effect sizes of g = 0.52 suggesting that most studies were underpowered to reliably detect medium-sized effects.*

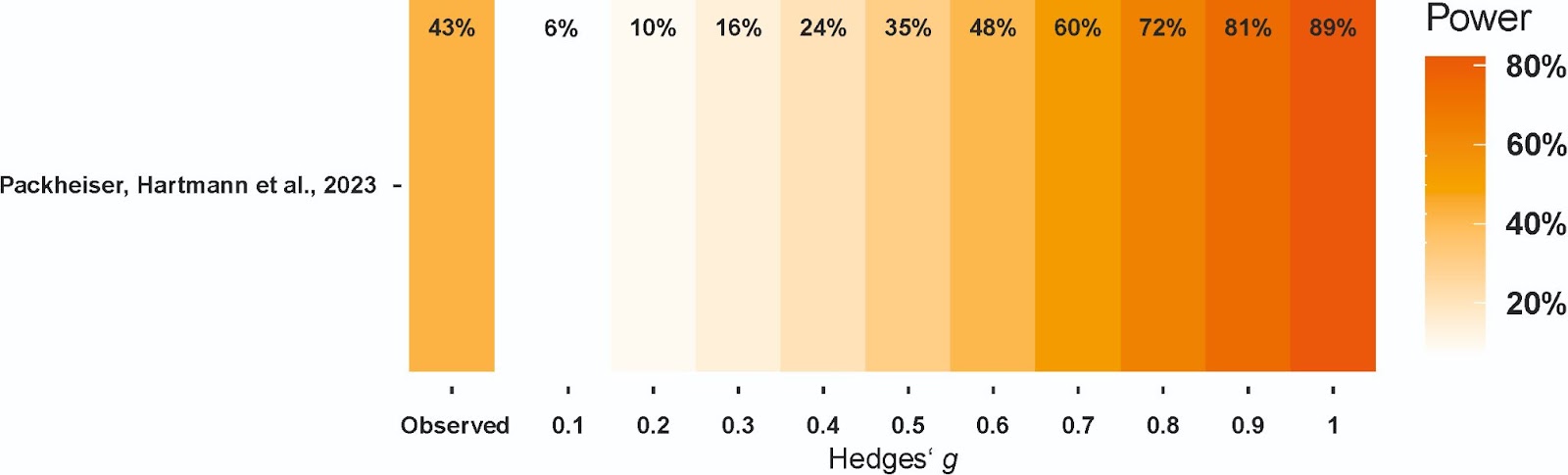

***Figure S3. Firepower plot (Quintana, 2022) for the meta-analysis of newborns studies.*** *The median power of each individual study in this meta-analysis was 43% to detect effect sizes of g = 0.54 suggesting that most studies were underpowered to reliably detect medium-sized effects.*

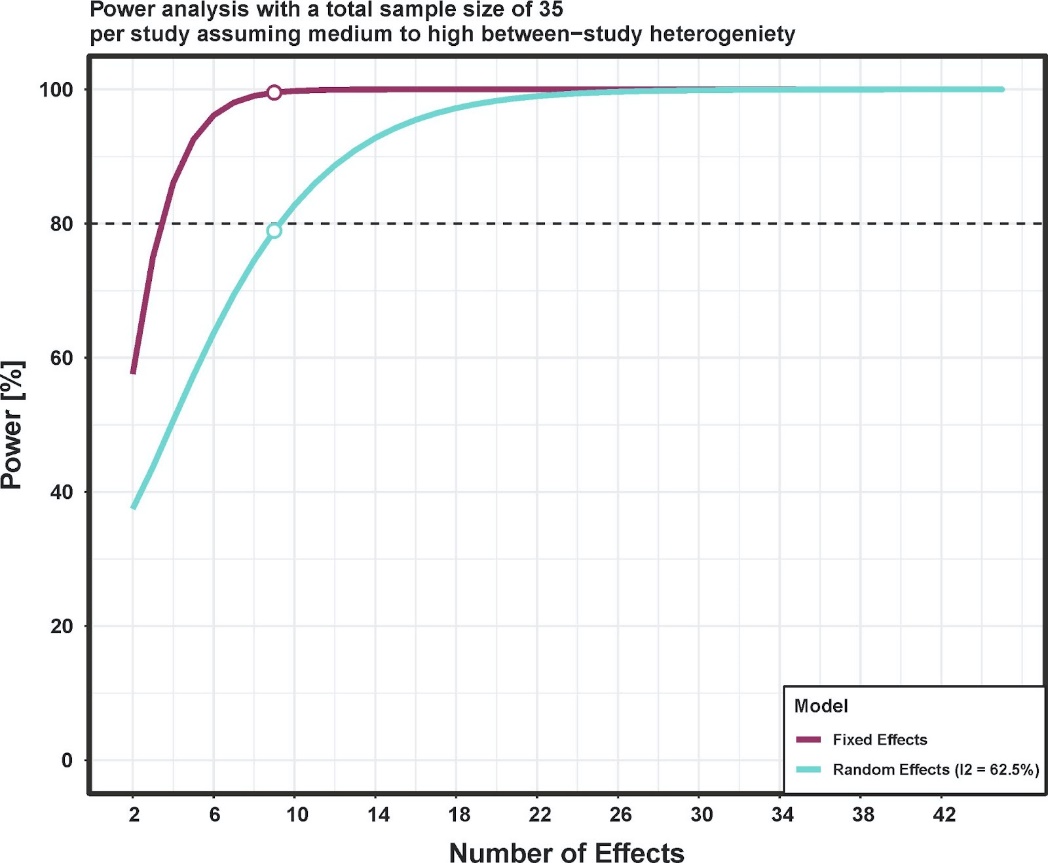

***Figure S4. Power calculations for the meta-analysis of children/adults studies using the mpower package (Nguyen et al., 2022).*** *Random effects models reached 80% power assuming medium to high heterogeneity at around nine effects.*

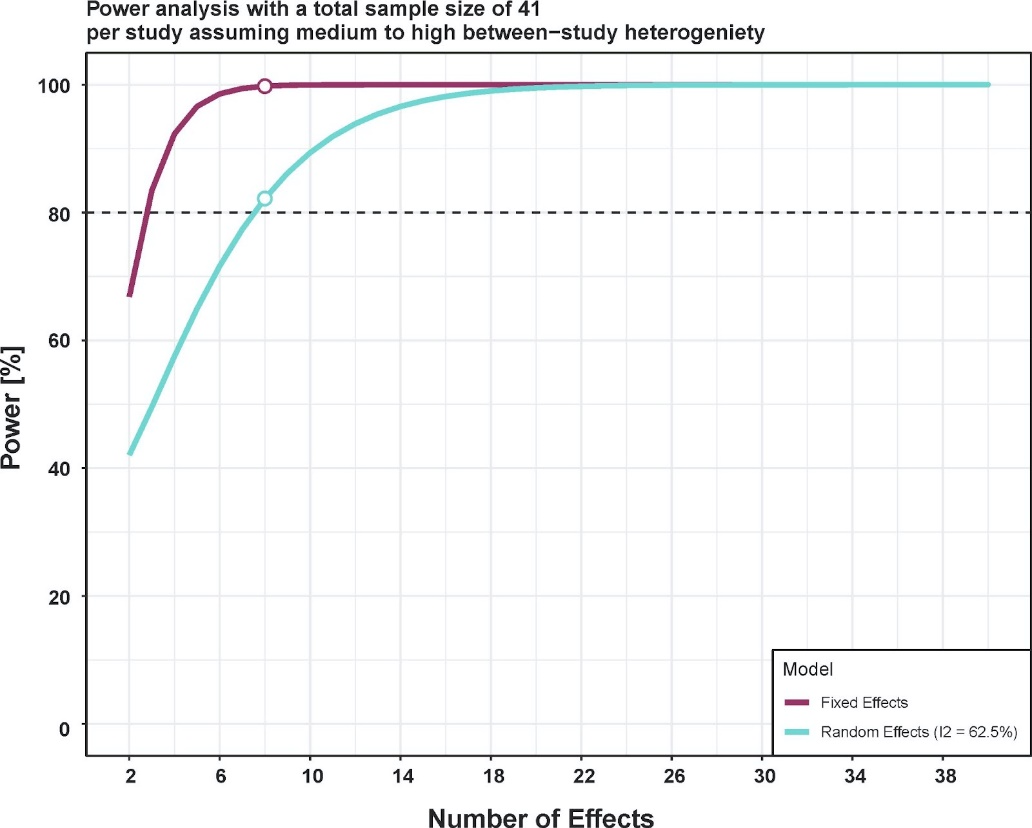

***Figure S5. Power calculations for the meta-analysis of newborns studies using the mpower package (Nguyen et al., 2022)..*** *Random effects models reached 80% power assuming medium to high heterogeneity at around nine effects.*

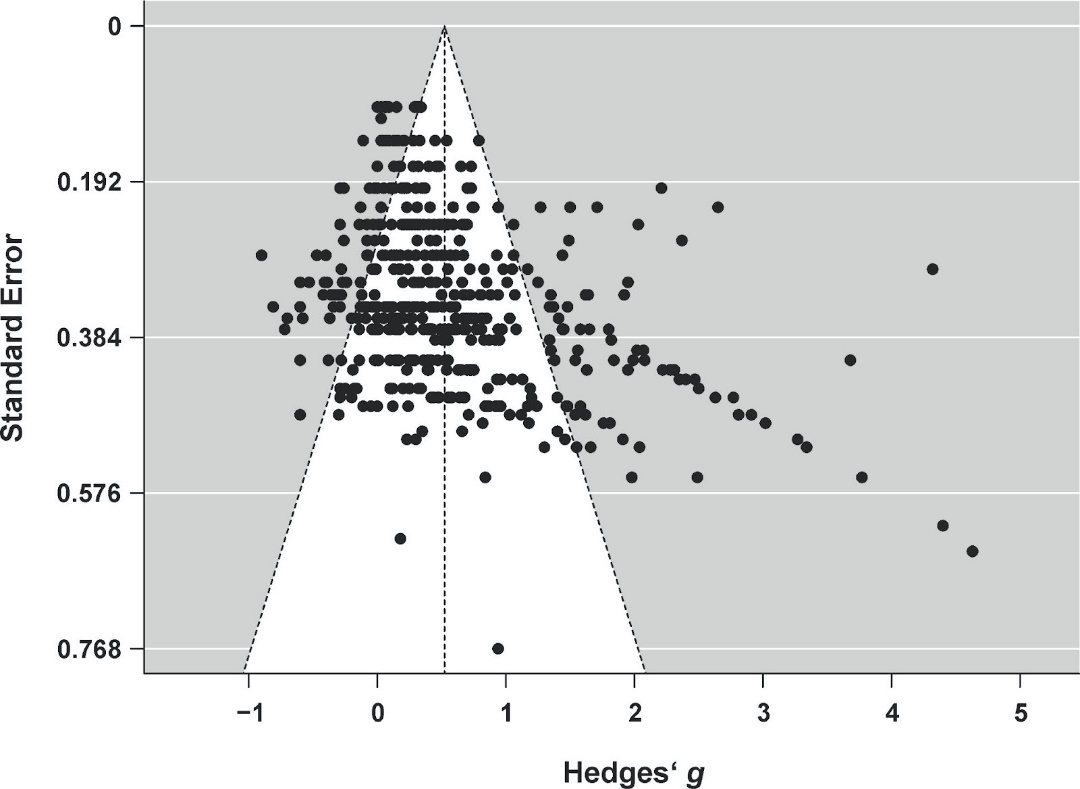

***Figure S6. Funnel plot for the adult meta-analysis.*** *Small study bias is indicated by funnel plot asymmetry.*

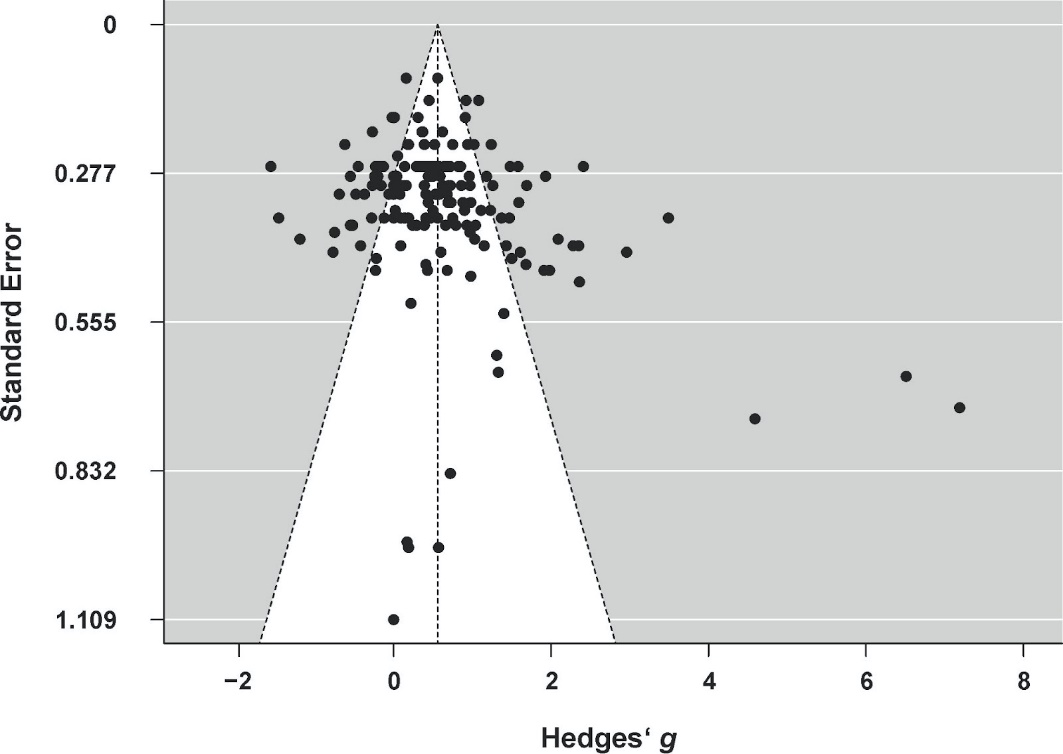

***Figure S7. Funnel plot for the meta-analysis of newborns studies.*** *No small study bias was detected.*

***Table S1. Alphabetized list of final outcomes separate for newborns and adults.*** *The table is separated by outcomes with sufficient power for further analysis and outcomes for which sufficient effects could not be extracted to allow for further analysis. Outcomes are colorized as physical health (PH) effects in magenta and mental health (MH) effects in blue. The number of available effects per outcome is listed in brackets. NA = not categorized.*

| **Outcomes for newborns that were analyzed at the specific outcome level  (*n* ≥ 8 effects)** | **Outcomes for adults that were analyzed at the specific outcome level (*n* ≥ 9 effects)** |
| --- | --- |
| Bowel movements (10, PH) | Anxiety, state (78, MH) |
| Cortisol (8, PH) | Anxiety, trait (25, MH) |
| Heart rate (12, PH) | Depression, trait (33, MH) |
| Liver enzymes (9, PH) | Diastolic/systolic blood pressure (21 each, PH) |
| Respiration (12, PH) | Fatigue (10, PH) |
| Temperature (9, PH) | Heart rate (31, PH) |
| Weight (45, PH) | Mobility (10, PH) |
|  | Negative affect (33, MH) |
|  | Pain (65, PH) |
|  | Positive affect (18, MH) |
|  | Respiration (13, PH) |
|  | Sleep (14, PH) |
| **Outcomes for newborns that were not analyzed at the specific outcome level  (*n* < 8 effects)** | **Outcomes for adults that were analyzed at the specific outcome level (*n* < 9 effects)** |
| Affect regulation (5, MH) | Affect regulation (1, MH) |
| Arousal (3, MH) | Anxiety, state/trait (3, MH) |
| Autonomic stability (3, PH) | Arousal (7, MH) |
| Bacterial decolonization (1, PH) | Autism behavior (1, MH) |
| Blood sugar (1, PH) | Balance (2, PH) |
| Death rate (3, PH) | Blood sugar (1, PH) |
| Depression, trait (1, MH) | Body satisfaction (1, MH) |
| Food intake (7, PH) | Disability (2, PH) |
| Grimacing (2, NA) | Disease assessment (1, PH) |
| Hospital stay (7, PH) | Emotional function (1, MH) |
| Immune system (2, PH) | Food intake (5, PH) |
| Motor function (4, PH) | Hospital stay (1, PH) |
| Negative affect, state (5, MH) | Immune system (5, PH) |
| Oxygen saturation (5, PH) | Insulin (1, PH) |
| Pain (3, PH) | Intellectual function (1, MH) |
| pH levels (1, PH) | Motor function (1, PH) |
| Postnatal complications (1, PH) | Muscle strength (1, PH) |
| Sleep (7, PH) | Nausea (4, PH) |
| Stress (7, MH) | Oxygen saturation (1, PH) |
| Urinary bone metabolism (1, PH) | Relaxation (6, MH) |
|  | Social function (3, MH) |
|  | Somatic/vegetative state (1, PH) |
|  | Somatization (1, PH) |
|  | Stereotypical behavior (1, MH) |
|  | Stress (7, MH) |
|  | Well-being, trait (5, MH) |

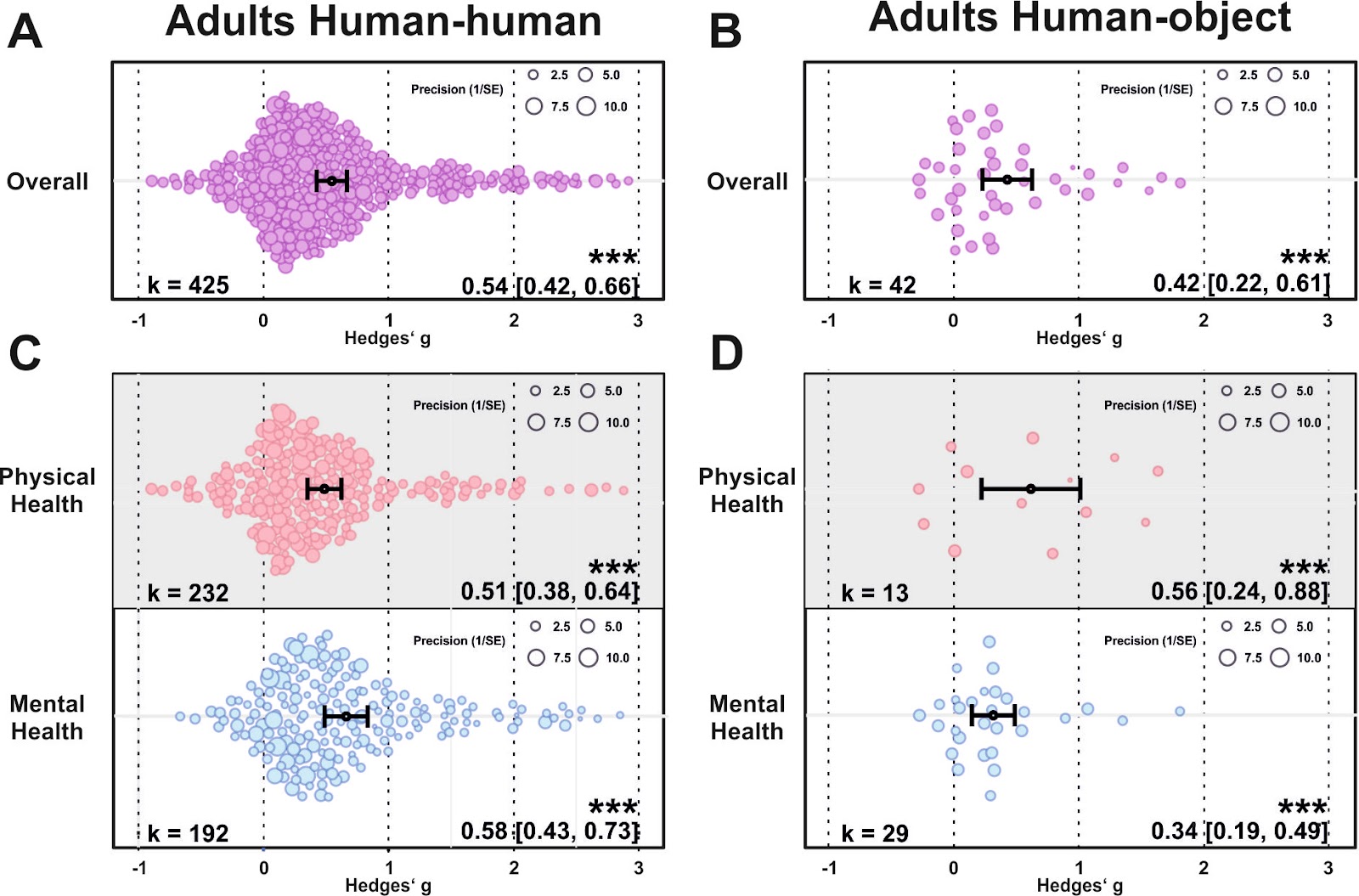

***Figure S8.*** (A) O*rchard plot illustrating the overall benefits across all health outcomes for human-human (A) and human-object touch (B). (C-D) same as (A-B) but separating the results for physical vs mental health benefits. Each dot reflects a measured effect and the number of effects (k) included in the analysis is depicted in the bottom left. Mean effects and 95% CIs are presented in the bottom right. Asterisks indicate the overall effect being significant from a null effect (*** p < .001, ** p < .01, * p < .05). Dot size reflects precision of each individual effect (larger = higher precision).*

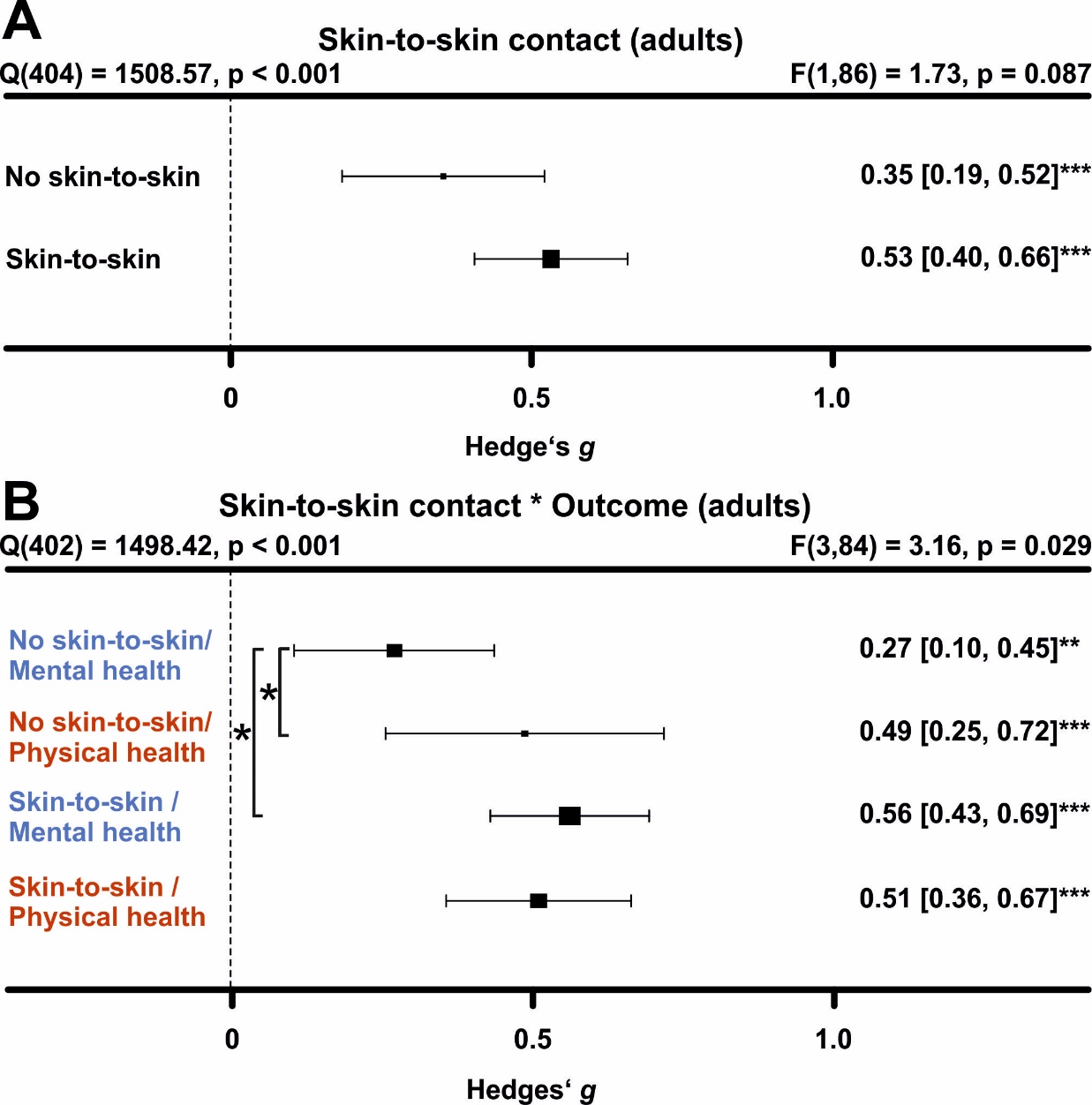

***Figure S9. Comparing health benefits of skin-to-skin contact during touch.*** *(A) Health benefits for present and absent skin-to-skin contact. All studies were included in this analysis, i.e. the analysis includes human-human, human-object and human-animal studies. All studies without human-human interactions were classified as absent skin-to-skin contact. (B) Same as A, but separating mental vs physical health benefits. Numbers on the right represent the mean effect, its 95% CI in square brackets and the significance level estimating the likelihood that the effect is equal to zero. The F-value in the top right represents a test of the hypothesis that all effects within the subpanel are equal. The Q statistic represents heterogeneity. Asterisks indicate the overall effect being significant from a null effect (*** p < .001, ** p < .01, * p < .05). Physical outcomes are marked in red, mental outcomes are marked in blue.*

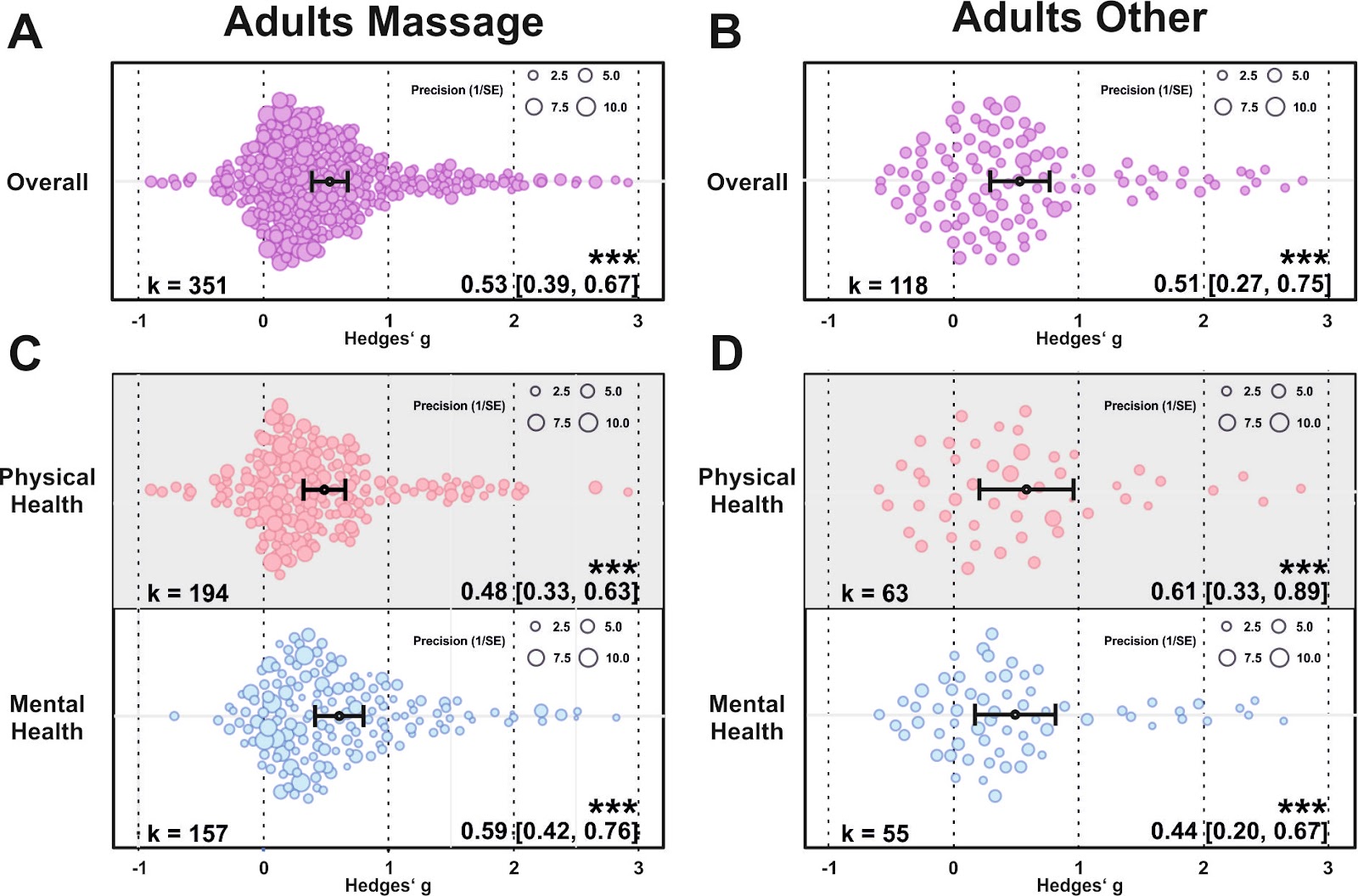

***Figure S10.*** *(A) Orchard plot illustrating the overall benefits across all health outcomes for massage interventions (A) and other types of touch interventions in adult cohorts (B). (C-D) same as (A-B) but separating the results for physical vs mental health benefits. Each dot reflects a measured effect and the number of effects (k) included in the analysis is depicted in the bottom left. Mean effects and 95% CIs are presented in the bottom right. Asterisks indicate the overall effect being significant from a null effect (*** p < .001, ** p < .01, * p < .05). Dot size reflects precision of each individual effect (larger = higher precision).*

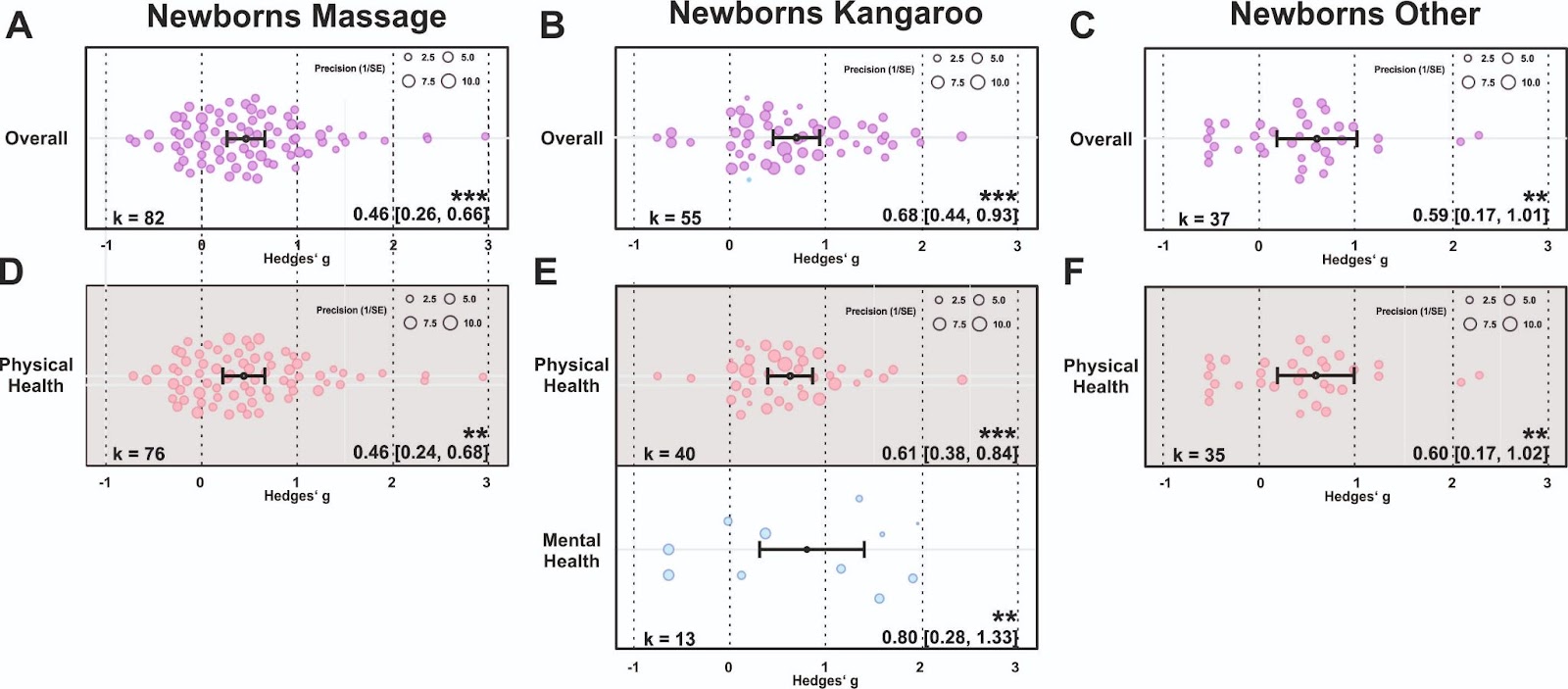
***Figure S11.*** *(A) Orchard plot illustrating the overall benefits across all health outcomes for massage interventions (A), the effects of kangaroo care (B) and other types of touch interventions in newborns (C). (E-F) same as (A-C) but separating the results for physical vs mental health benefits if sufficient effects for further analysis could be found. Each dot reflects a measured effect and the number of effects (k) included in the analysis is depicted in the bottom left. Mean effects and 95% CIs are presented in the bottom right. Asterisks indicate the overall effect being significant from a null effect (*** p < .001, ** p < .01, * p < .05). Dot size reflects precision of each individual effect (larger = higher precision).*

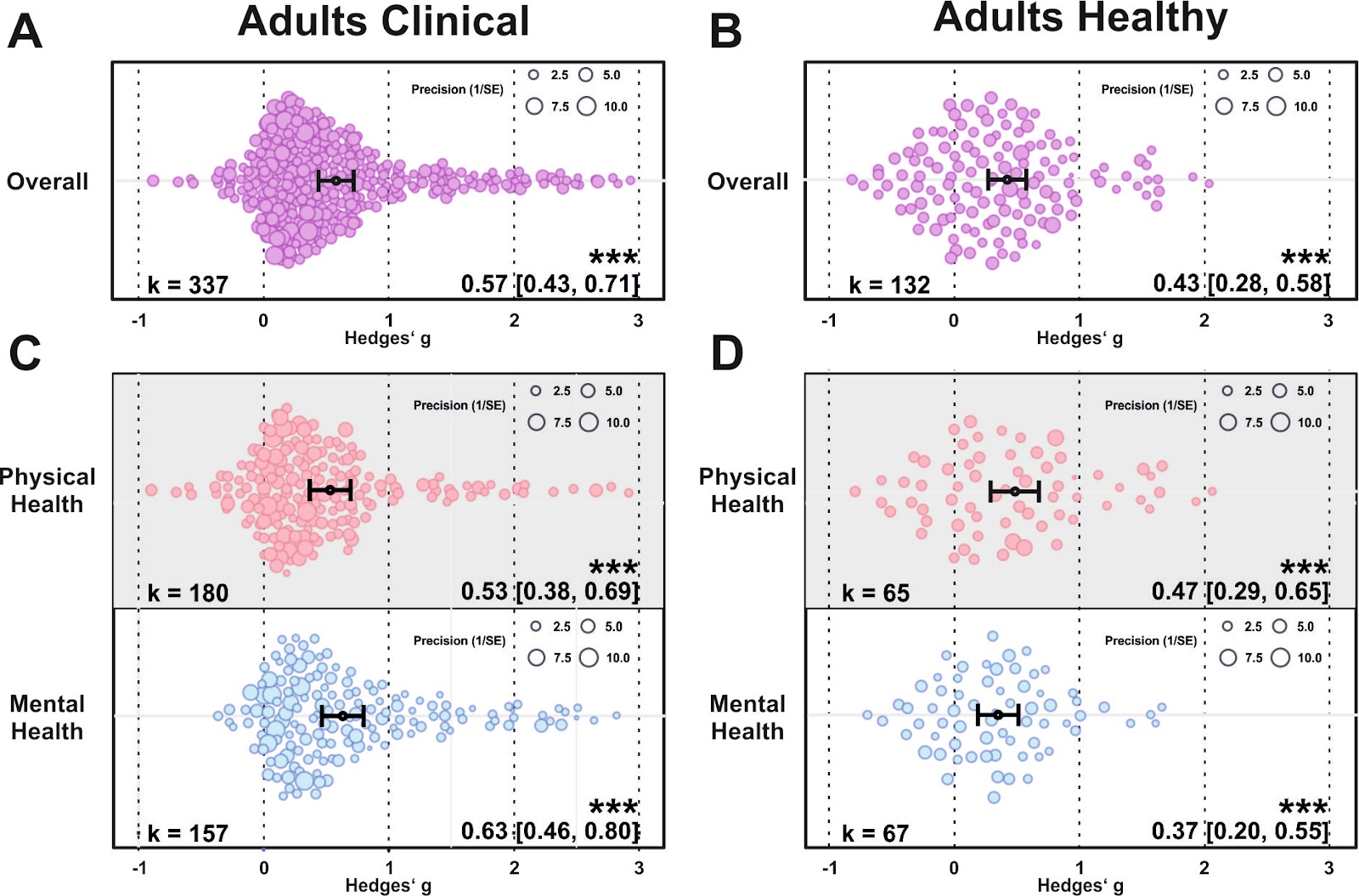

***Figure S12.*** *(A) Orchard plot illustrating the overall benefits across all health outcomes for clinical (A) and healthy adult cohorts (B). (C-D) same as (A-B) but separating the results for physical vs mental health benefits. Each dot reflects a measured effect and the number of effects (k) included in the analysis is depicted in the bottom left. Mean effects and 95% CIs are presented in the bottom right. Asterisks indicate the overall effect being significant from a null effect (*** p < .001, ** p < .01, * p < .05). Dot size reflects precision of each individual effect (larger = higher precision).*

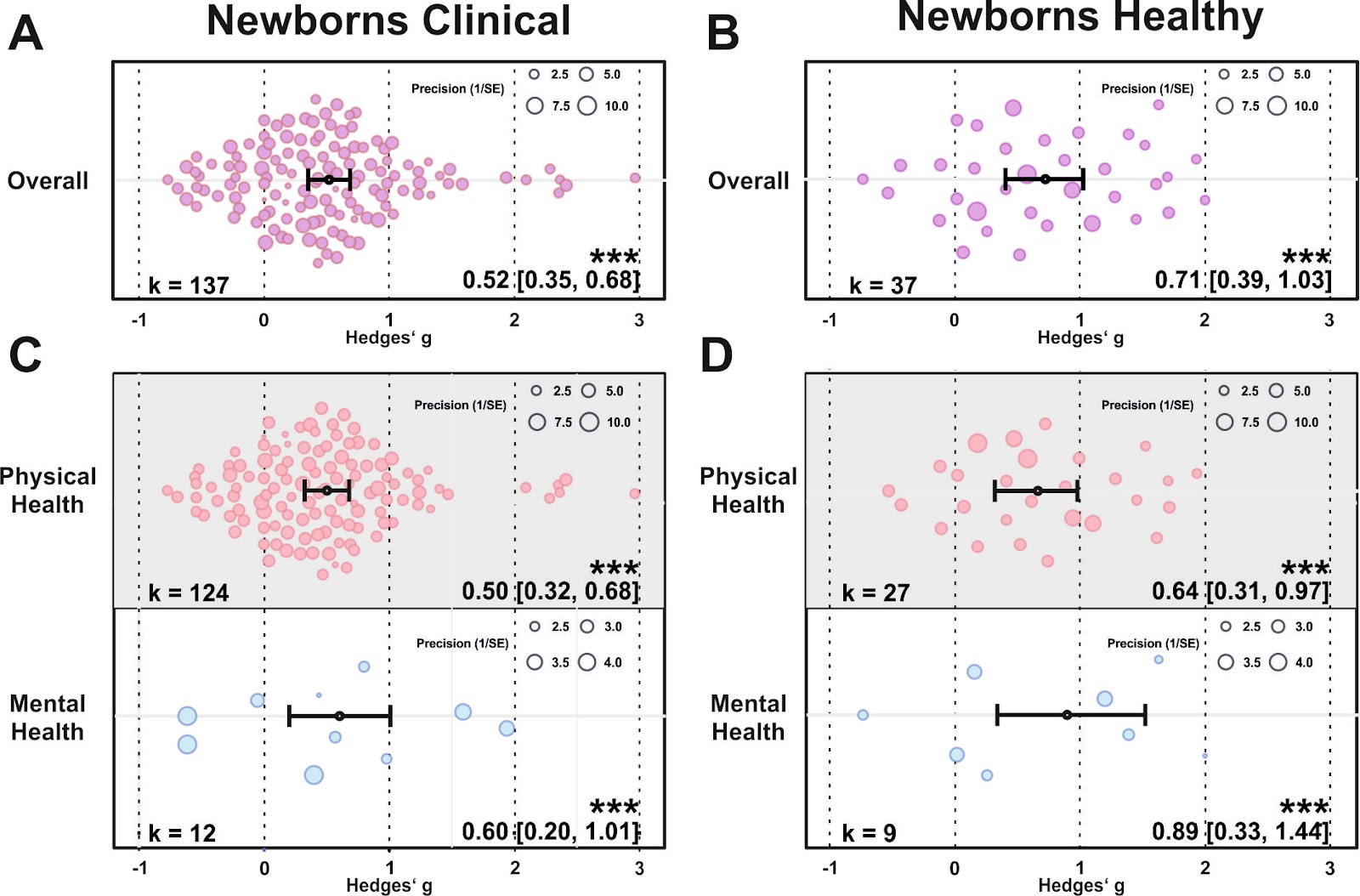

***Figure S13.*** *(A) Orchard plot illustrating the overall benefits across all health outcomes for clinical (A) and healthy newborns (B). (C-D) same as (A-B) but separating the results for physical vs mental health benefits. Each dot reflects a measured effect and the number of effects (k) included in the analysis is depicted in the bottom left. Mean effects and 95% CIs are presented in the bottom right. Asterisks indicate the overall effect being significant from a null effect (*** p < .001, ** p < .01, * p < .05). Dot size reflects precision of each individual effect (larger = higher precision).*

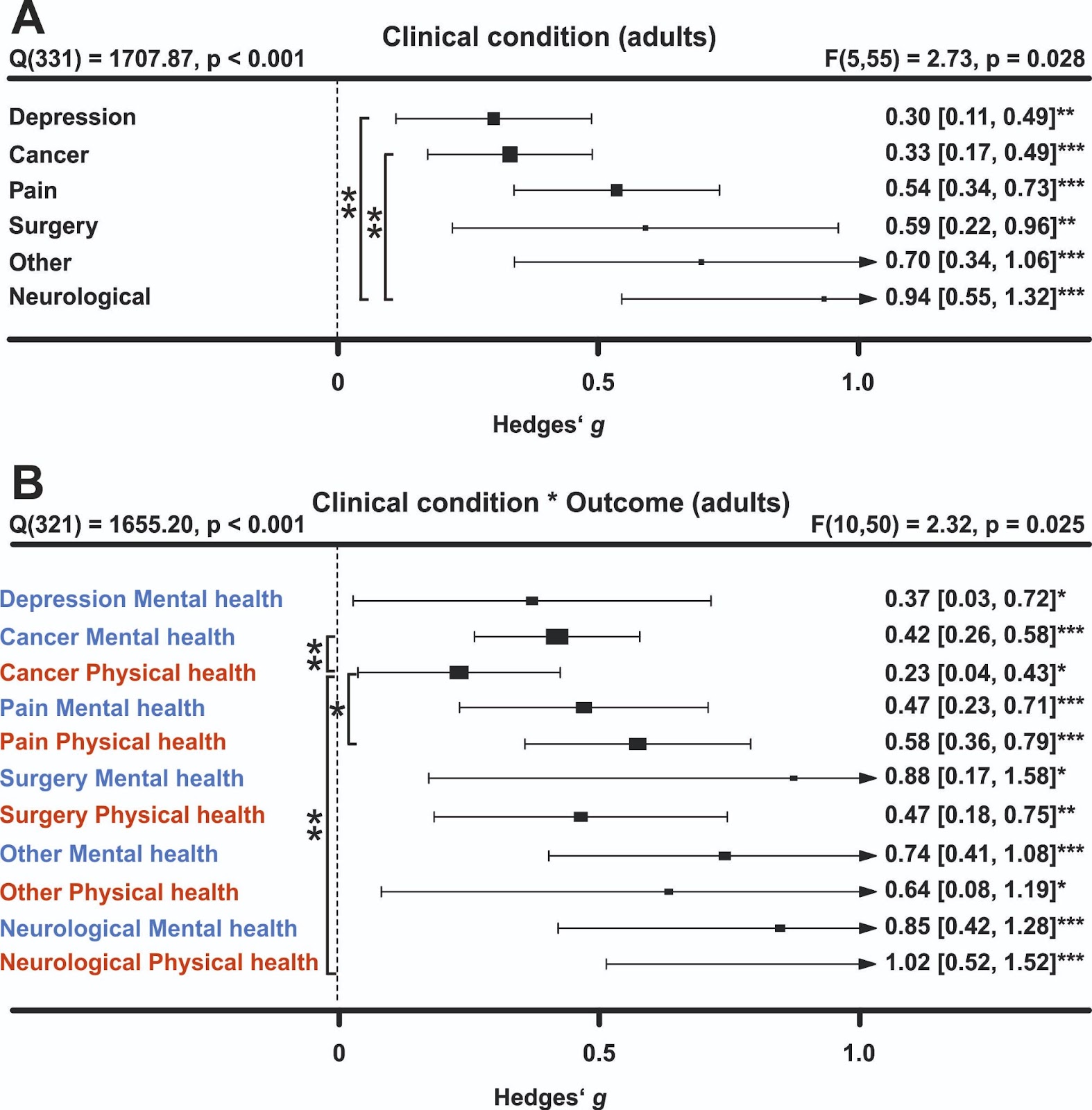

***Figure S14. Comparing health benefits across different clinical disorders.*** *(A) Health benefits in individuals diagnosed with depression, cancer, pain syndromes, following surgery, neurological or other types of disorders. (B) Same as A, but separating mental vs physical health benefits. Numbers on the right represent the mean effect, its 95% CI in square brackets and the significance level estimating the likelihood that the effect is equal to zero. The F-value in the top right represents a test of the hypothesis that all effects within the subpanel are equal. The Q statistic represents heterogeneity. Asterisks indicate the overall effect being significant from a null effect (*** p < .001, ** p < .01, * p < .05). Physical outcomes are marked in red, mental outcomes are marked in blue.*

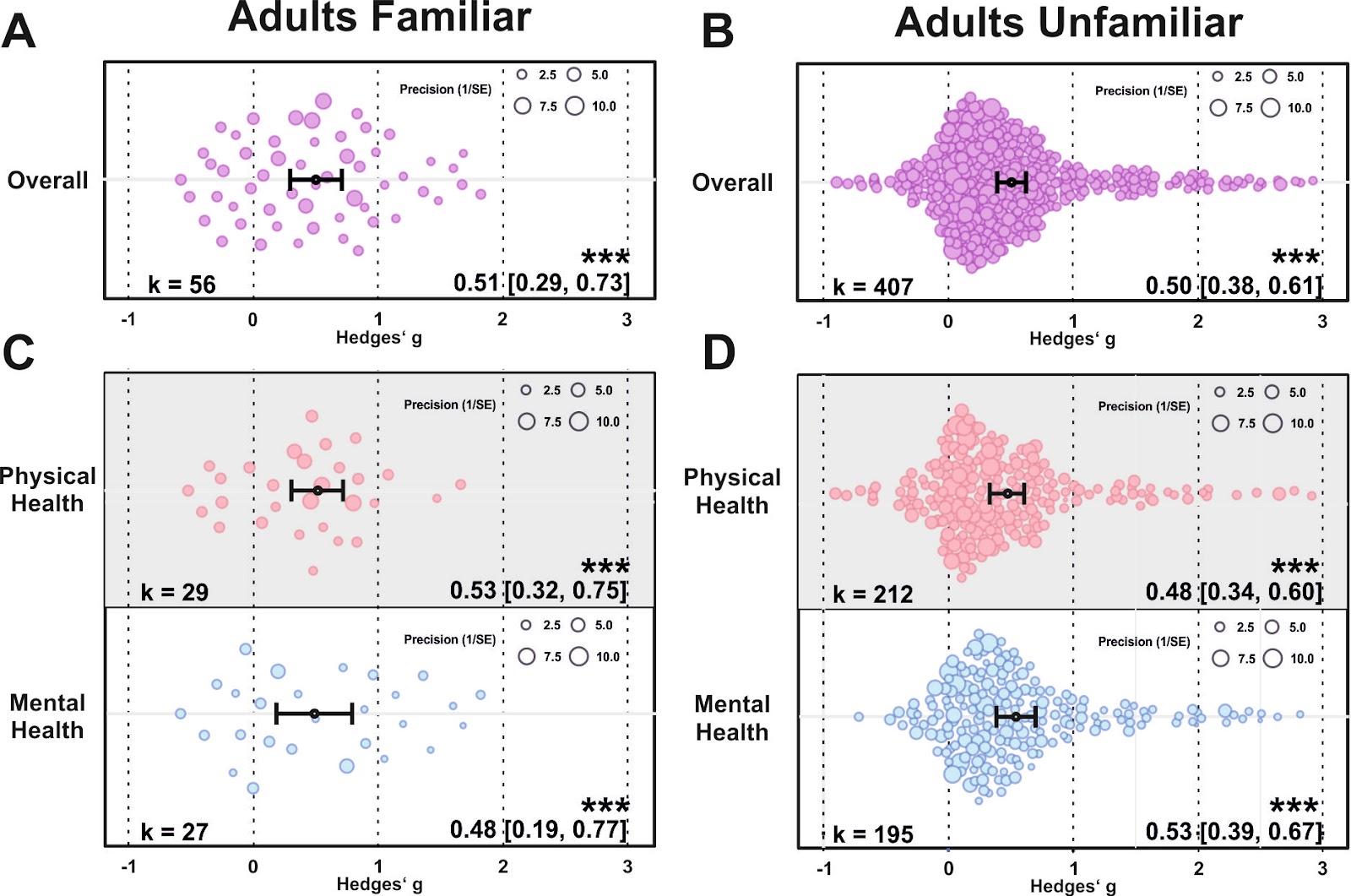

***Figure S15.*** *(A) Orchard plot illustrating the overall benefits across all health outcomes for touch applied by a familiar (A) and unfamiliar individual in adult cohorts (B). (C-D) same as (A-B) but separating the results for physical vs mental health benefits. Each dot reflects a measured effect and the number of effects (k) included in the analysis is depicted in the bottom left. Mean effects and 95% CIs are presented in the bottom right. Asterisks indicate the overall effect being significant from a null effect (*** p < .001, ** p < .01, * p < .05). Dot size reflects precision of each individual effect (larger = higher precision).*

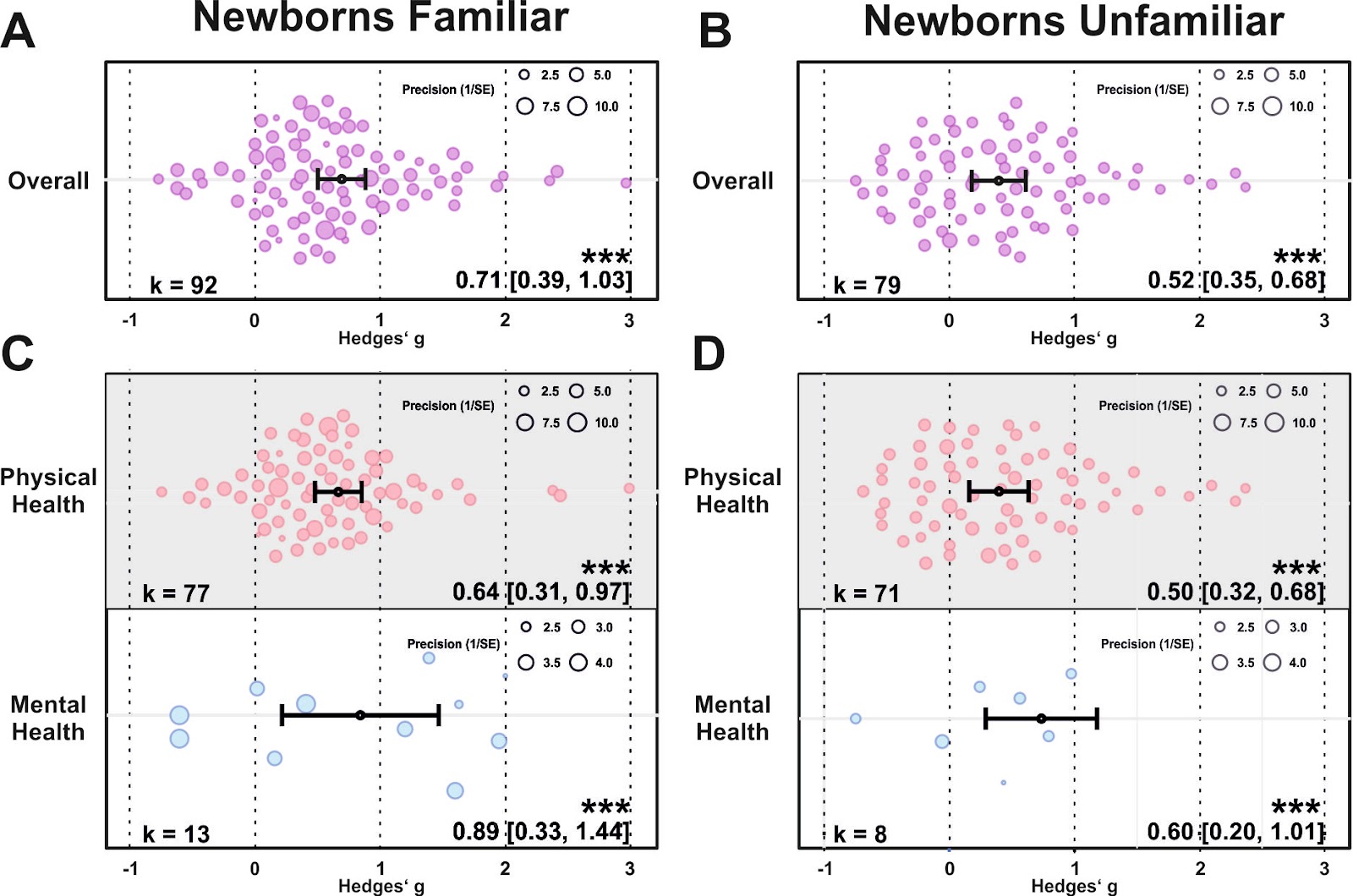

***Figure S16.*** *(A) Orchard plot illustrating the overall benefits across all health outcomes for touch applied by a familiar (A) and unfamiliar individual in newborns (B). (C-D) same as (A-B) but separating the results for physical vs mental health benefits. Each dot reflects a measured effect and the number of effects (k) included in the analysis is depicted in the bottom left. Mean effects and 95% CIs are presented in the bottom right. Asterisks indicate the overall effect being significant from a null effect (*** p < .001, ** p < .01, * p < .05). Dot size reflects precision of each individual effect (larger = higher precision).*

*Body part*

For the touched body part, we found significantly higher health benefits for head touch compared to arm touch (*p* = .039) and torso touch (*p* = .031; see Supplementary Figure S17). Touching the arm resulted in lower mental health compared to physical health benefits (*p* = .028). Furthermore, we found a significantly increased physical health benefit when the head was touched as opposed to touch of the torso (*p* = .043). Thus, head touch such as a face or scalp massage could be especially beneficial.

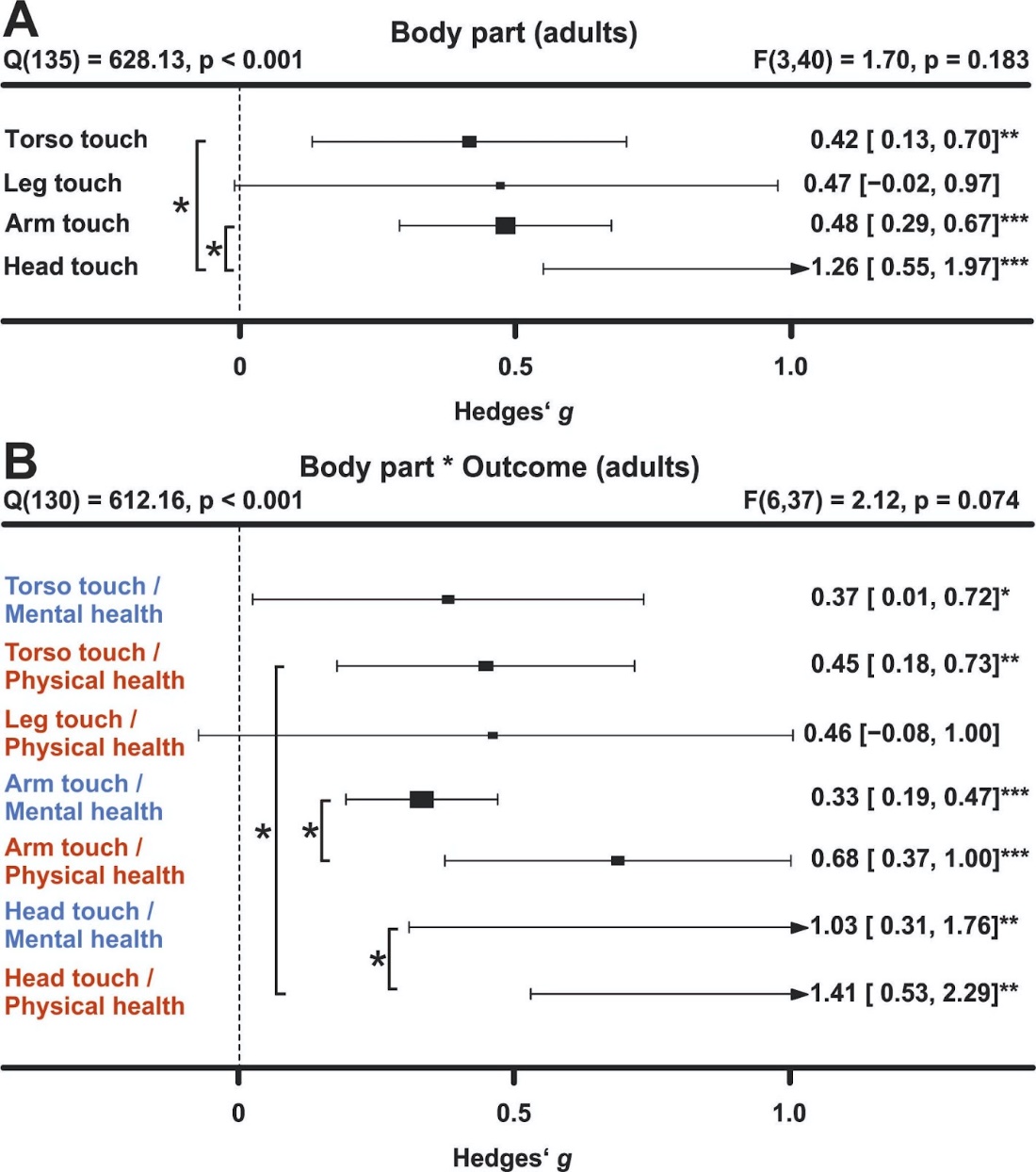

***Figure S17. Comparing health benefits depending on the touched body part.*** *(A) Health benefits depend on the location where touch was applied (arm, leg, torso or head). Studies in which multiple body parts were touched were excluded from the analysis. (B) Same as A, but separating mental vs physical health benefits. Note that there was an insufficient number of effects for further analysis of mental health benefits following leg touch. Asterisks indicate the overall effect being significant from a null effect (*** p < .001, ** p < .01, * p < .05).*

*Familiarity of touch location*

Familiarity with the location in which the touch was applied (familiar being for example the participants’ home) did not influence the efficacy of touch interventions (Supplementary Figure S18).

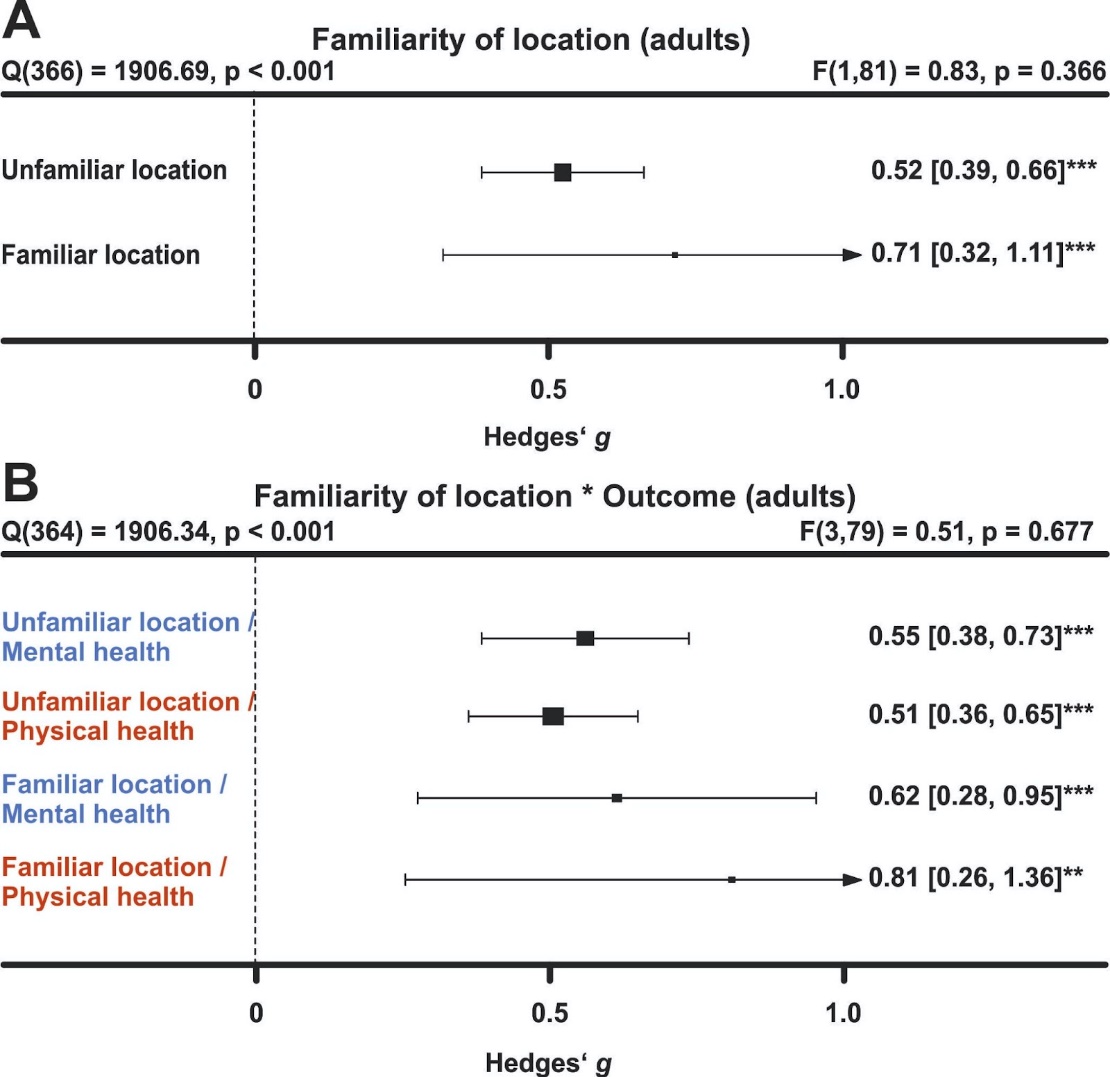

***Figure S18. Comparing health benefits depending on familiarity with the  intervention location.*** *(A) Health benefits separated by intervention location in adults (A) and newborns (B). (C-D) Same as (A-B), but separating mental vs physical health benefits. Numbers on the right represent the mean effect, its 95% CI in square brackets and the significance level estimating the likelihood that the effect is equal to zero. The F-value in the top right represents a test of the hypothesis that all effects within the subpanel are equal. The Q statistic represents heterogeneity. Asterisks indicate the overall effect being significant from a null effect (*** p < .001, ** p < .01, * p < .05). Physical outcomes are marked in red, mental outcomes are marked in blue.*

*Directionality*

In adults, we tested whether a uni- or bi-directional application of touch mattered. The large majority of touch was applied unidirectionally (*k* = 442 of 469 effects). Unidirectional touch had higher health benefits (*p* = .032). Specifically mental health benefits were higher in unidirectional touch (*p* = .045).

*Study location*

For adults, we found significantly stronger health benefits of touch in South American compared to North American cohorts (*p* = .026) and European cohorts (*p* = .029). For newborns, we found stronger effects in Asian and European cohorts compared to North American cohorts (both ps < .05). Investigating the interaction with mental and physical health benefits did not reveal any effects of study location in both meta-analyses (see Supplementary Figure S19).

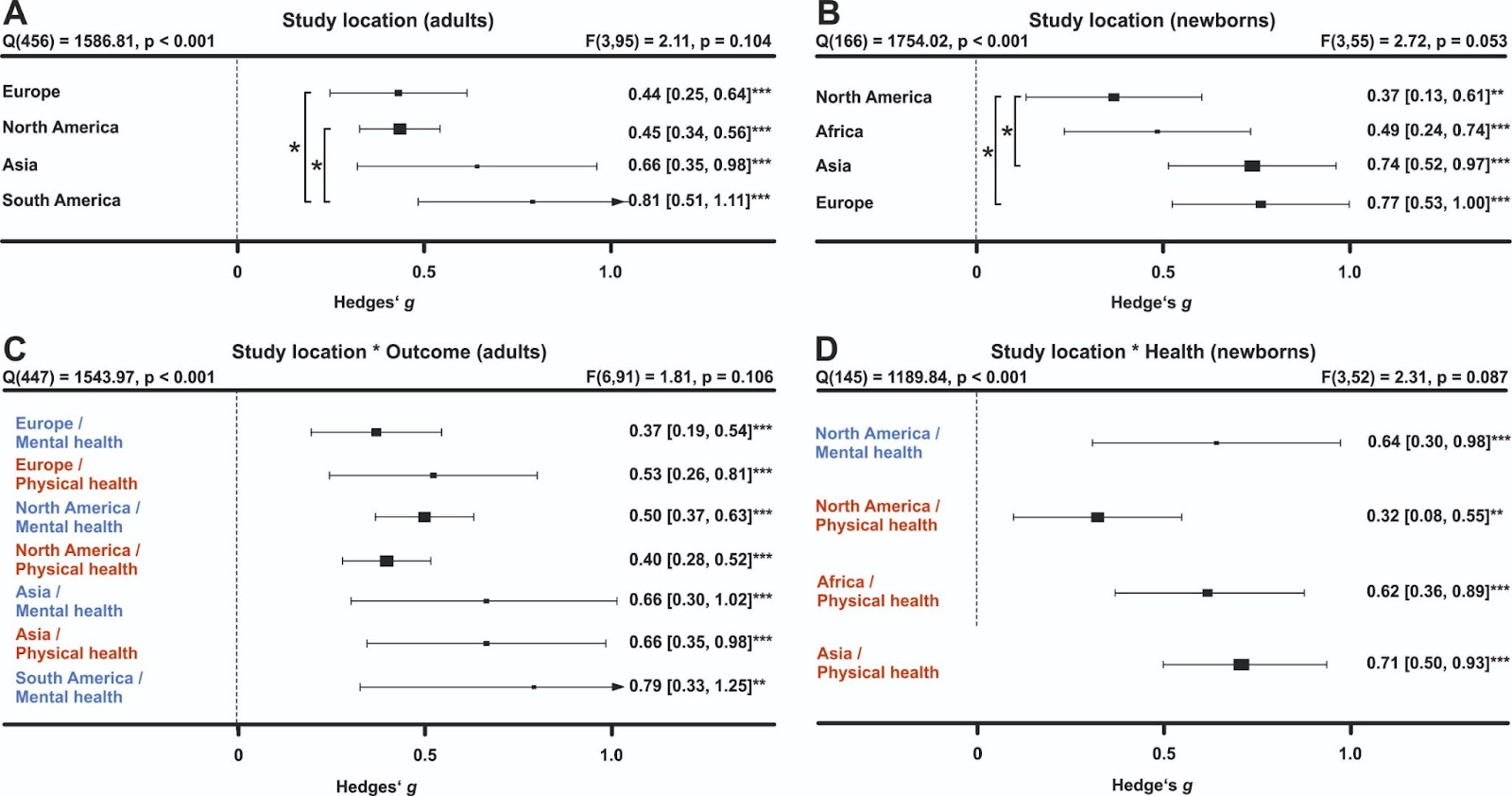

***Figure S19. Comparing health benefits depending on study location.*** *(A) Health benefits separated by study location in adults (A) and newborns (B). (C-D) Same as (A-B), but separating mental vs physical health benefits. Note that there was an insufficient number of effects for further analysis for certain regions both for the overall and the specific effects on physical or mental health in both meta-analyses. Numbers on the right represent the mean effect, its 95% CI in square brackets and the significance level estimating the likelihood that the effect is equal to zero. The F-value in the top right represents a test of the hypothesis that all effects within the subpanel are equal. The Q statistic represents heterogeneity. Asterisks indicate the overall effect being significant from a null effect (*** p < .001, ** p < .01, * p < .05). Physical outcomes are marked in red, mental outcomes are marked in blue.*

*Systematic review*

All studies where data could not be obtained or that did not meet the meta-analysis inclusion criteria can be found [here](https://osf.io/c8rvw/?view_only=978a5dace61f4a43806b2d39895844d3) in the file “Study_lists_final.xlsx” (sheet “Studies_without_effect_sizes”). For human health outcomes assessed across *k* = 56 studies and *n* = 2438 individuals, 90.0% of mental health and 84.3% of physical health parameters were positively impacted by touch for datasets where no effect size could be computed. This matches well with the observations of the meta-analysis of a highly positive benefit of touch overall.

As part of our systematic review, we also assessed health outcomes in animals across *k* = 19 studies and *n* = 911 subjects. For animal studies, 71.4% of effects showed benefits to mental health-like parameters and 81.8% showed positive physical health effects. We thus found strong evidence that touch interventions, which were mostly conducted by humans, had positive health effects in animal species as well. More specifically, beneficial effects of touch in animals were comparably strong for mental health-like and physical health outcomes. This may inform interventions to promote animal welfare in the context of animal experiments (Lewejohann et al., 2020), farming (Sørensen et al., 2001) and pets (Verga & Michelazzi, 2009). While most studies investigated effects in rodents mostly used as laboratory animals, these results likely transfer to livestock and common pets as well: touch was beneficial in lambs, fish and cats (Coulon et al., 2015; Soares et al., 2011; Gourkow et al., 2014). The positive impact of human touch in rodents also allows for future mechanistic studies in animal models to investigate how interventions such as tickling or stroking modulate hormonal and neuronal responses to touch in the brain. Furthermore, the commonly proposed oxytocin hypothesis can be causally investigated in these animal models through for example optogenetic or chemogenetic techniques (Oliveira et al., 2021). We believe that such translational approaches will further help in optimizing future interventions in humans by uncovering the underlying mechanisms and brain circuits involved in touch.
